# SCARF: Auto-Segmentation Clinical Acceptability & Reproducibility Framework for Benchmarking Essential Radiation Therapy Targets in Head and Neck Cancer

**DOI:** 10.1101/2022.01.15.22269276

**Authors:** Joseph Marsilla, Jun Won Kim, Denis Tkachuck, Sejin Kim, Joshua Siraj, John Cho, Ezra Hahn, Ali Hosni, Kristine Jacinto, Mattea L. Welch, Michal Kazmierski, Katrina Rey-McIntyre, Shao Hui Huang, Tirth Patel, Tony Tadic, Fei-Fei Liu, Scott Bratman, Andrew Hope, Benjamin Haibe-Kains

## Abstract

**Background and Purpose:** Auto-segmentation of organs at risk (OAR) in cancer patients is essential for enhancing radiotherapy planning efficacy and reducing inter-observer variability. Deep learning auto-segmentation models have shown promise, but their lack of transparency and reproducibility hinders their generalizability and clinical acceptability, limiting their use in clinical settings.

**Materials and Methods:** This study introduces SCARF (auto-Segmentation Clinical Acceptability & Reproducibility Framework), a comprehensive six-stage reproducible framework designed to benchmark open-source convolutional neural networks for auto-segmentation of 19 essential OARs in head and neck cancer (HNC).

**Results:** SCARF offers an easily implementable framework for designing and reproducibly benchmarking auto-segmentation tools, along with thorough expert assessment capabilities. Expert assessment labelled 16/19 AI-generated OAR categories as acceptable with minor revisions. Boundary distance metrics, such as 95th Percentile Hausdorff Distance (95HD), were found to be 2x more correlated to Mean Acceptability Rating (MAR) than volumetric overlap metrics (DICE).

**Conclusions:** The introduction of SCARF, our auto-Segmentation Clinical Acceptability & Reproducibility Framework, represents a significant step forward in systematically assessing the performance of AI models for auto-segmentation in radiation therapy planning. By providing a comprehensive and reproducible framework, SCARF facilitates benchmarking and expert assessment of AI-driven auto-segmentation tools, addressing the need for transparency and reproducibility in this domain. The robust foundation laid by SCARF enables the progression towards the creation of usable AI tools in the field of radiation therapy. Through its emphasis on clinical acceptability and expert assessment, SCARF fosters the integration of AI models into clinical environments, paving the way for more randomised clinical trials to evaluate their real-world impact.

Highlights

- Our study highlights the significance of both quantitative and qualitative controls for benchmarking new auto-segmentation systems effectively, promoting a more robust evaluation process of AI tools.
- We address the lack of baseline models for medical image segmentation benchmarking by presenting SCARF, a comprehensive and reproducible six-stage framework, which serves as a valuable resource for advancing auto-segmentation research and contributing to the foundation of AI tools in radiation therapy planning.
- SCARF enables benchmarking of 11 open-source convolutional neural networks (CNN) against 19 essential organs-at-risk (OARs) for radiation therapy in head and neck cancer, fostering transparency and facilitating external validation.
- To accurately assess the performance of auto-segmentation models, we introduce a clinical assessment toolkit based on the open-source QUANNOTATE platform, further promoting the use of external validation tools and expert assessment.
- Our study emphasises the importance of clinical acceptability testing and advocates its integration into developing validated AI tools for radiation therapy planning and beyond, bridging the gap between AI research and clinical practice.

## Introduction

In recent years, deep-learning based architectures have dominated the field of automated segmentation of organs at risk (OAR) in the head and neck region [1–13]. However, the lack of transparency in publishing auto-segmentation methods, particularly regarding the release of code and data used for model training, has been a persistent issue [14]. While some studies have demonstrated the potential of deep learning-based auto-segmentation (DLAS) methods for optimising clinical contouring workflows, there remains a gap in understanding whether predicted contours gain clinician approval [8,12]. Some studies have placed an emphasis on clinical assessment of contours produced by these auto segmentation models without providing insight into whether the predicted contours actually receive clinician approval. Studies publishing auto-segmentation solutions across a wide range of medical image segmentation tasks have a tendency to disregard both reproducibility of their methods and the subsequent evaluation of their methods in a clinical setting to validate their findings. There have been a large number of guidelines and other papers describing requirements for publishing sufficient information to assess model performance [15,16]. However, these guidelines rarely provide a ready-made infrastructure or systematic code-base to realise the intent of the guidelines.

To address these challenges and promote both reproducibility and clinical evaluation of segmentation methods, we developed the auto-Segmentation Clinical Acceptability & Reproducibility Framework (SCARF). This six-step framework empowers researchers and clinicians to build and robustly evaluate open-source networks for various segmentation tasks related to radiation therapy and beyond. SCARF’s steps can be easily adapted to improve existing open-source segmentation repositories and checklists focused on leveraging artificial intelligence modelling for clinical applications [15,17].

Emphasising transparency through reproducibility of data, code, and tools, SCARF provides an intuitive framework for packaging research outputs, ensuring transparency, reproducibility, and reusability of medical imaging models. Moreover, it offers a collection of published auto-segmentation models and their corresponding training and evaluation protocols, facilitating robust benchmarking of baseline performance for medical segmentation tasks. To address the critical aspect of clinical acceptability, SCARF leverages the open-source QUANNOTATE platform [18], enabling internal and external evaluation of segmentation methods by clinical experts.

We demonstrate the application of SCARF in the development of deep learning models for the delineation of 19 essential OARs in head and neck cancer radiation therapy. In summary, SCARF, with its compendium of open-source and reproducible auto-segmentation models, curated datasets, and clinical acceptability testing toolkit, lays a strong foundation for the advancement and benchmarking of the next generation of deep learning models in medical imaging segmentation.

## Materials and Methods

### Dataset Curation

Radiological and clinical data extracted from institutional databases or public repositories, such as The Cancer Imaging Archive (TCIA) [19], often requires a high level of curation to harmonise the data and make them ready for deep learning. In this study, we used a large institutional imaging dataset of 3211 HNC patients whose radiological and clinical data were available (https://doi.org/10.7937/J47W-NM11) [20] . A UHN institutional review board approved our study (REB 17-5871); we performed all experiments in accordance with relevant guidelines and ethical regulations of Princess Margaret Cancer Centre (PM). The associated clinical data have been collected prospectively as part of the PM Anthology of Outcomes [21]. We used Med-ImageTools [22] to collect the meta-data and curate the radiation therapy structure files (RT-STRUCT).

### Curation of Auto-Segmentation Models

We have identified a set of auto-segmentation models that have been published between 2016 and 2020 for which sufficient data, code and documentation have been released to allow re-implementation. A literature search was conducted from January 2016 to April 2020 to find medical image segmentation studies using deep learning-based modelling approaches. We reviewed 75 studies with a medical image segmentation theme in the conducted literary search to augment networks we could test during our analysis. We refactored all tested models into a Pytorch Lightning framework [23] to increase readability, reusability and shareability of the re-implementation code.

### Training of Auto-Segmentation Models

Models were trained using a combined loss scheme to address the heavy pixel-wise class imbalance present within our dataset between the individual classes of OARs. For each experiment we used the same 80%/10%/10% split, corresponding to 479, 44, and 59 scans for training, tuning, and testing, respectively. Each model was trained on 4 NVIDIA Tesla P100 GPUs for 3 days or until convergence. Early stopping was implemented if no significant change (0.1 decrease in loss magnitude) was made in tuning loss minimization after 50 epochs. More information regarding specifics in configuring our training pipeline can be found in Appendix A.

### Performance Evaluation

The performance of each model was estimated by averaging volumetric overlap indices (Dice similarity coefficient - DICE, jaccard index) with boundary distance metric (95th% Hausdorff Distance - 95HD) on the independent testing set of patients (**Supplementary Figure 1**) [24].Taking this into consideration, after fine tuning the winning model in a second round of training, four other quantitative performance metrics were calculated. Additional metrics calculated include boundary metrics (Surface Distance [SD], Added path length [APL]), and false negative metrics (False Negative Volume and False Negative Length) [25]. Multiple metrics are essential during validation of any segmentation task as overlap-based metrics such as DICE do not take the correctness (or complexity) of an object’s boundaries into account. Additionally, properties of the target structure need to be considered when evaluating the scores for each OAR. For example, a small pixel deviation in a low volume OAR like the chiasm, can have a substantial effect on DICE and other volumetric based overlap metrics that are calculated for it [26]. More information regarding model finetuning and selection can be found in Appendix A.

### Clinical Evaluation

We employed an enhanced QUANNOTATE interface to facilitate the review and evaluation of clinical acceptability by multiple physician observers. They rated AI-generated and Ground Truth OAR contour pairs while remaining blinded to the source of origin for each contour. (**Supplementary Figure 2**) [24]. Mean Acceptability Rating (MAR) was calculated for each contour examined by averaging ratings across all observers. After inference, segmentation network performance was assessed by calculating six different performance metrics listed in **Performance Evaluation**. These metrics were extracted for each OAR contour. To determine the weight each metric should have when assessing acceptability, we computed the correlation between each performance metric and MAR using Pearson to identify which metrics best align with clinical acceptability [27–29]. Overlap metrics were considered ‘more clinically acceptable’ if they showed significantly positive correlation with MAR, while boundary distance metrics were considered ‘more clinically acceptable’ if they showed significantly negative correlation with MAR. Suggestions will be made as to how these metrics can be used to optimise a network’s segmentations for clinical acceptability.

### Statistical Analysis

To evaluate the statistical power of our study, a continuous endpoint, two independent sample study power analysis was conducted for each OAR group. The MAR from each of the 4 observers can be averaged and separated into two distinct study groups for each OAR category. (Group A: MAR of Ground-Truth contours; Group B: MAR of AI-Generated contours). Because no analysis has been conducted to this effect for each OAR category previously, the MAR and standard deviation for each OAR category were taken from the final calculated MAR after clinical acceptability testing was completed. Post-hoc power (PHP) analysis was also conducted for each OAR category. We will use these preliminary acceptability testing results to further refine sample size in future clinical acceptability testing experiments.

### Generalizability Assessment

Our model was trained using RADCURE as the primary dataset. Due to the inevitable biases intrinsic to demographics of patients treated at our centre, we tested whether our model performed accurately on data collected at external institutions with varying patient populations. A model’s generalizability, in this context, is the ability of a model to perform as trained when applied to external data collected at different institutions. To assess model generalizability, seven publically available datasets were collected and curated (Supplementary Table 1).

### Data Availability

Raw imaging data and corresponding contours are available on The Cancer Imaging Archive (TCIA) [19] (Supplementary Table 1). Processed external datasets and predictions using our model on those datasets are publicly available via the ptl-oar-segmentation GitHub repository (https://github.com/bhklab/ptl-oar-segmentation).

### Code Availability

The computer code for the reimplementation and evaluation of all the auto-segmentation models reimplemented in this study is available via https://github.com/bhklab/ptl-oar-segmentation. The code for the QUANNOTATE clinical acceptability testing interface is available via the Quannotate GitHub repository (https://github.com/bhklab/quannotate).

### Model Availability

The weights for each trained auto segmentation model trained, and processed versions of each external dataset, were saved and made available on the project’s github page. All auto segmentation models have been uploaded to mhub.ai to maximise reusability.

### Research Reproducibility

Tutorials were generated for easy re-implementation using <u>Google Collab.</u> Users can use these collaborative notebooks provided to get started with easy re-implementation. Users can also clone the github repositories, set-up a local anaconda environment based on the one provided, set their local variables and train each network using internal resources. Templates have also been provided to integrate their own networks and training protocols if desired.

## Results

### Auto-Segmentation Clinical Acceptability & Reproducibility Framework (SCARF)

To improve the development of auto-segmentation models and their potential clinical impact, we propose a framework composed of six main steps: (1) Dataset Curation, (2) Model Selection, (3) Model Training, (4) Quantitative Performance Evaluation, (5) Clinical Assessment, and (6) Generalizability Assessment. (Figure 1). Successful implementation of this new framework requires a reproducible way to extract and curate medical imaging data and metadata. Furthermore, providing a reproducible architecture for selection, training and evaluation of open-source models can help the community curate “baseline” data for a wide range of segmentation tasks. Finally, placing a strong emphasis on clinical acceptance, and providing open-source tools to conduct these assessments will help optimise these systems for increased clinical benefit, which translates to greater adoption and use of these models by clinicians at large. SCARF’s goal is to build a community resource that allows collection, standardisation, testing and validation of various segmentation methods made available by the open-source community with the intent of establishing baseline quantitative and qualitative data for each region of interest being segmented (**Figure 1**).

**Figure 1:**
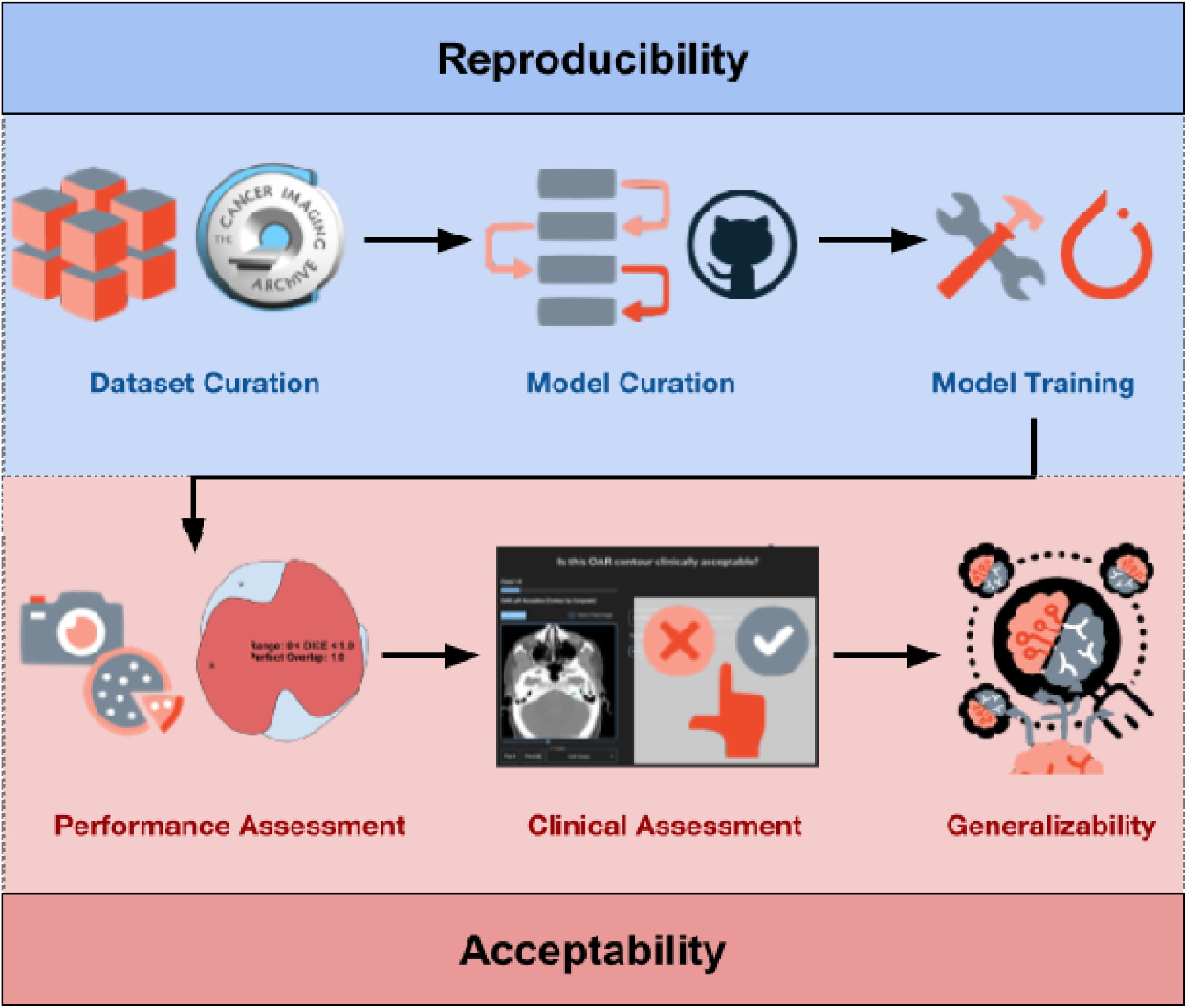
SCARF Overview: auto-Segmentation Clinical Acceptability & Reproducibility Framework. Reproducibility: The first half of the cycle emphasizes reproducible development of code and software used during dataset curation, model selection and model training stages. Acceptability: In the second half of the cycle, emphasis is placed on development of an acceptability standard, that uses “clinician focused” evaluation protocols to identify relevant performance metrics and build quality assurance tools that focus on recording data that will aid model optimization to maximize expert approval ratings.

### Dataset Curation

SCARF provides an open-source methodology to rapid dataset curation, model benchmarking and clinical performance assessment for radiation oncology specific segmentation tasks. For this analysis, we selected 19 OARs that were consistently delineated in a subset of 582 head and neck cancer patients in RADCURE (Supplementary Figure 3). Seven external datasets, spanning a total of 587 patients [12,55–59], were also collected and curated to assess the generalizability of the best auto-segmentation models with variability of overlapping OARs (Supplementary Tables 1 and 2).

### Model Curation & Training

SCARF’s codebase enables rapid integration and training of open-source models coded in Pytorch to benchmark these networks on your segmentation task. We selected 11 open-source CNNs to train on the segmentation of the 19 OARs of interest [7,30–39] (**Supplementary Figure 4, Supplementary Table 3**).

### Performance Evaluation

SCARF enables the comparison and performance validation using multiple metrics which are necessary for accurately representing model performance. For initial performance evaluation, we use volumetric overlap metrics (DICE) and boundary distance metrics, which assess the error at the boundary of two overlapping contours (95HD, SD) for the combined set of OARs being segmented [5,10,13]. When analysing the mean performance metrics of all OARs for each open source model trained, the top three segmentation models were UNET variants. WOLNET (a simple implementation of the standard 3D-UNET) [40,41] performed best among the 11 models and achieved the highest average DICE (0.765±0.10) (**Figure 2A**) and lowest average HD (2.63±2.61) (**Figure 2B**). Altered 3D versions of UNet3+DEEPSUP [34,42] (DICE 0.74±0.10; HD 2.98±2.63) and UNet++ [35,42] (DICE 0.73±0.10; HD 3.10±2.83) were the second and third top-performing segmentation models, respectively (**Supplementary Figure 5**).

**Figure 2:**
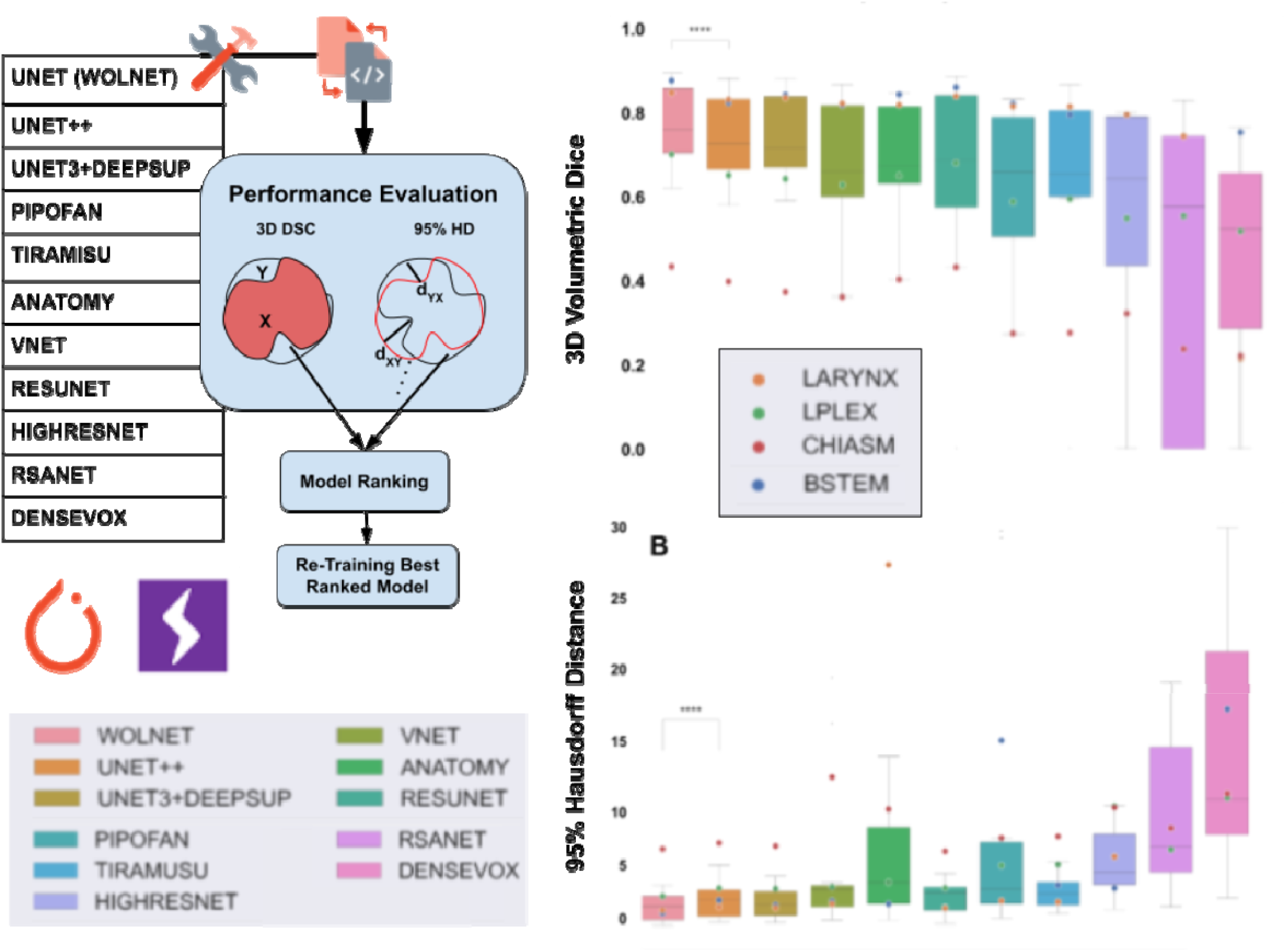
Performance Evaluation Of Model Training. Eleven open-source networks were selected based on availability, complexity and adaptability. Model(s) were each trained with 523 patient scans for 3 days on 4xTesla P100 GPU(s) or until convergence using PyTorch-lighting. Barplots of classical metrics (DICE and 95HD) plotted for all OAR(s) for each model when applied to a hold out set of 59 scans. Average values of Brainstem (BSTEM), Larynx, L Brachial-Plexus, and Optic Chiasm are plotted for each model. One simple 3D UNET architecture performed best across all OAR(s) segmented for both metrics. This network (labeled WOLNET, after its author) was chosen for retraining and use in clinical acceptability testing.

### Performance Evaluation of the Best Model After Fine Tuning

SCARF allows for optimisation and versioning of networks, which make tracking improvements made by hyperparameter or architecture changes easy. The training scheme of the best performing open source network, WOLNET, was further tuned resulting in a final average test DICE of (0.77± 0.09) and a 95HD of (3.42± 4.05) across all OAR(s). (Supplementary Figure 6 A-C).

### Clinical Evaluation

In addition to quantitative benchmarking of segmentation methods, SCARF implements an open-source web-based toolkit for rating clinical acceptability of contours generated for each region of interest being automatically delineated. In our experiment, four experienced oncologists from our centre used the QUANNOTATE interface to complete the blinded questionnaire defined above for the “Ground Truth” (human) and “AI-Generated” sets of contours for each OAR. When comparing MAR for all OARs, 78% of Ground-Truth contours are considered acceptable (**Figure 3A**) compared with 52% for AI-generated contours (**Figure 3B**). When analysing individual OAR categories, Ground-Truth contours were considered more acceptable than AI-generated contours for 15 out of the 19 OARs assessed. Experts rated 16/19 AI-Generated OARs as acceptable for planning with minor edits (3 < MAR < 3.5) (**Table 1**). Only three OAR categories (brainstem, larynx, and the right optic nerve) were shown to be rated more clinically acceptable than their paired deep learning contour with sufficient post-hoc power (PHP > 80%). Ten OAR categories had no significant differences in MAR (PHP < 20%) indicating that the WOLNET network can currently delineate these OARs with human level accuracy. The MAR between the remaining six OAR categories (20% < PHP < 0.8%) may be significantly different if more samples are analysed for each group (**Table 1**). Mean acceptability rating correlation with 6 common segmentation metrics was extracted. Mean acceptability rating showed significant negative correlation with boundary distance metrics like 95HD and Surface distances. (∼-0.26 for 95HD and ∼-0.30 for Surface Distance). A less significant positive correlation with DICE was also observed (∼0.14) (Appendix B, Supplementary Figure 7).

**Table 1:** MC v. DLC Acceptability Ratings For Each OAR Category. Pn80 is defined as the minimum power required (number of paired samples) to have a significantly different rating between Ground-Truth and AI contours. For example, for our analysis of the sample size When assessing whether certain OAR categories passed the mean acceptability cutoff of 3.5, 15 manually delineated OARs on average were considered clinically acceptable, requiring no edits for planning purposes, compared with 9 OARs generated by deep learning. When analyzing categories of OARs requiring minor edits for their contours to be accepted into radiation therapy plans (3.0 < MAR < 3.5), 7 deep learning generated OARs compared with 4 manually contoured OARs met this criteria.

| ROI | MAR (GT) | MAR (AI) | Pn80 | PHP % | ROI | MAR (GT) | MAR (AI) | Pn80 | PHP % |
| --- | --- | --- | --- | --- | --- | --- | --- | --- | --- |
| MAND | **4.15±0.89** | 3.53±1.13 | 32 | 27.5 |  |  |  |  |  |
| LEYE | **3.45±0.71** | 3.00±0.78 | 39 | 27.1 | RLENS | 4.10±0.78 | **4.15±0.70** | 3820 | 3.5 |
| REYE | **3.68±0.62** | 3.05±0.68 | 15 | 58.1 | LACOU | **3.30±0.91** | 3.20±0.97 | 1300 | 4.3 |
| BSTEM | **4.13±0.69** | 2.8±0.91 | **4** | **95.8** | RACOU | 3.30±0.72 | **3.38±0.74** | 1272 | 4.3 |
| LARYNX | **3.95±0.85** | 2.38±0.90 | **5** | **98** | LPLEX | 3.68±0.76 | **3.78±0.58** | 907 | 5.2 |
| SPCOR | **3.93±0.76** | 3.70±0.56 | 171 | 11.7 | RPLEX | **3.63±0.84\*** | 3.58±0.81 | 4431 | 3.4 |
| LPAR | 3.80±0.99 | **3.85±1.02** | 6154 | 3.2 | LIPS | **3.75±0.71** | 3.33±0.80 | 45 | 23.6 |
| RPAR | **4.08±1.02\*** | 3.95±1.23 | 966 | 4.4 | LOPTIC | **4.28±0.68** | 3.50±0.78 | 12 | 66.47 |
| ESOPH | **3.93±0.80** | 3.13±0.97 | 16 | 52.1 | ROPTIC | **4.18±0.68** | 3.20±0.79 | **8** | **84.5** |
| LLENS | **4.05±0.68** | 3.65±0.92 | 45 | 19.6 | CHIASM | **3.03±1.27\*** | 2.90±1.06 | 1498 | 4.3 |

**Figure 3:**
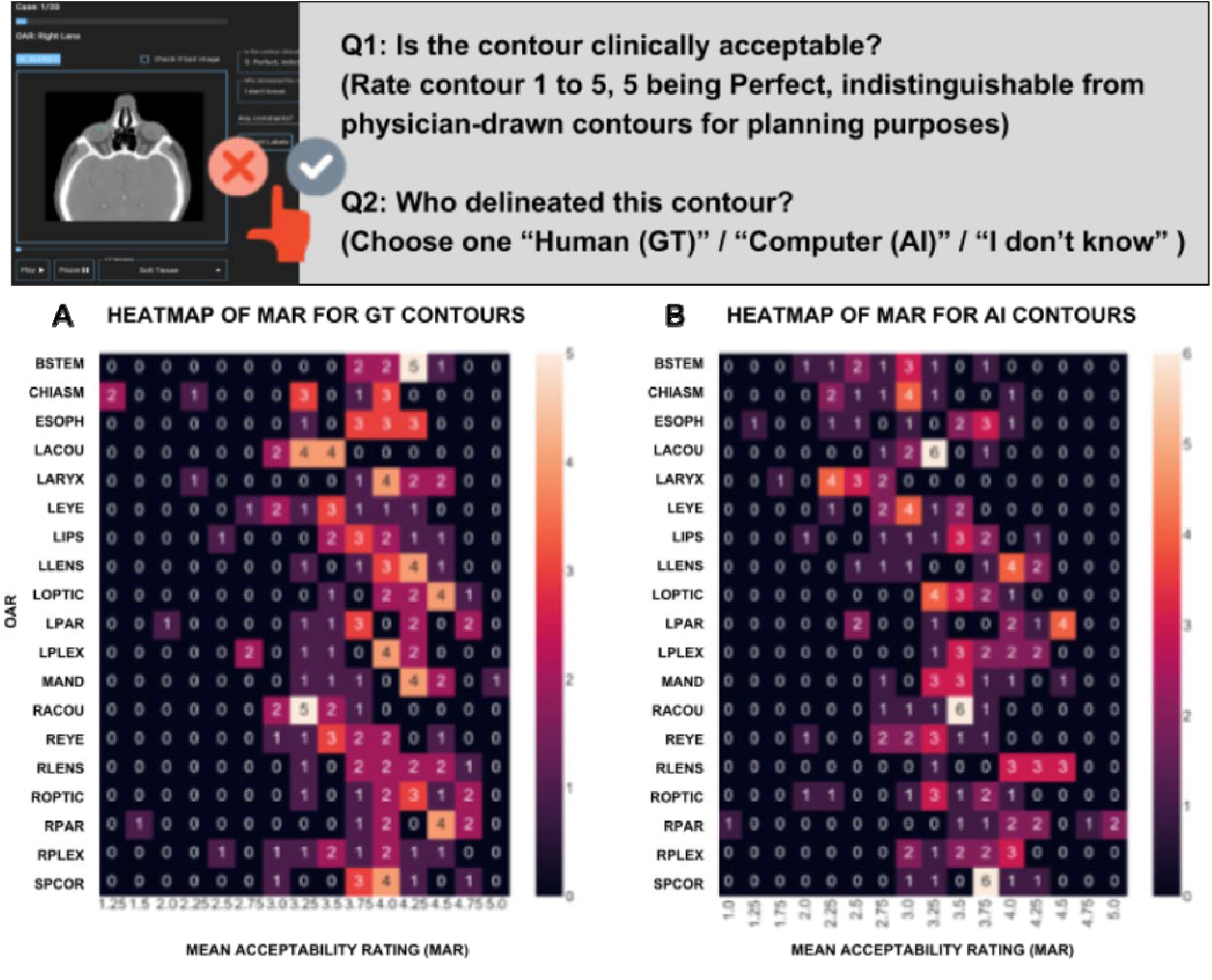
Results Of Clinical Evaluation Recording Using Quannotate QA Tool. Results of the acceptability test by representing mean acceptability rating (MAR) counts for each OAR in a heat-map for (**A**) Ground-Truth contours (GT) and (**B**) AI-generate contours (AI). The higher the value of a box the more contours of that given OAR (row) had any given MAR value (column) and the lighter that box will be. Notice a shift to the left when examining the heat-map of mean acceptability ratings for deep learning contours examined for each OAR indicating a greater degree of clinical acceptance for manual contours as depicted by figure 4D. GT contours received a significantly higher mean rating of 3.75 than AI contours which were rated 3.23 when all OARs were considered (3.75 ± 0.77 vs. 3.23 ± 0.86, p <0.01).

**Figure 4:**
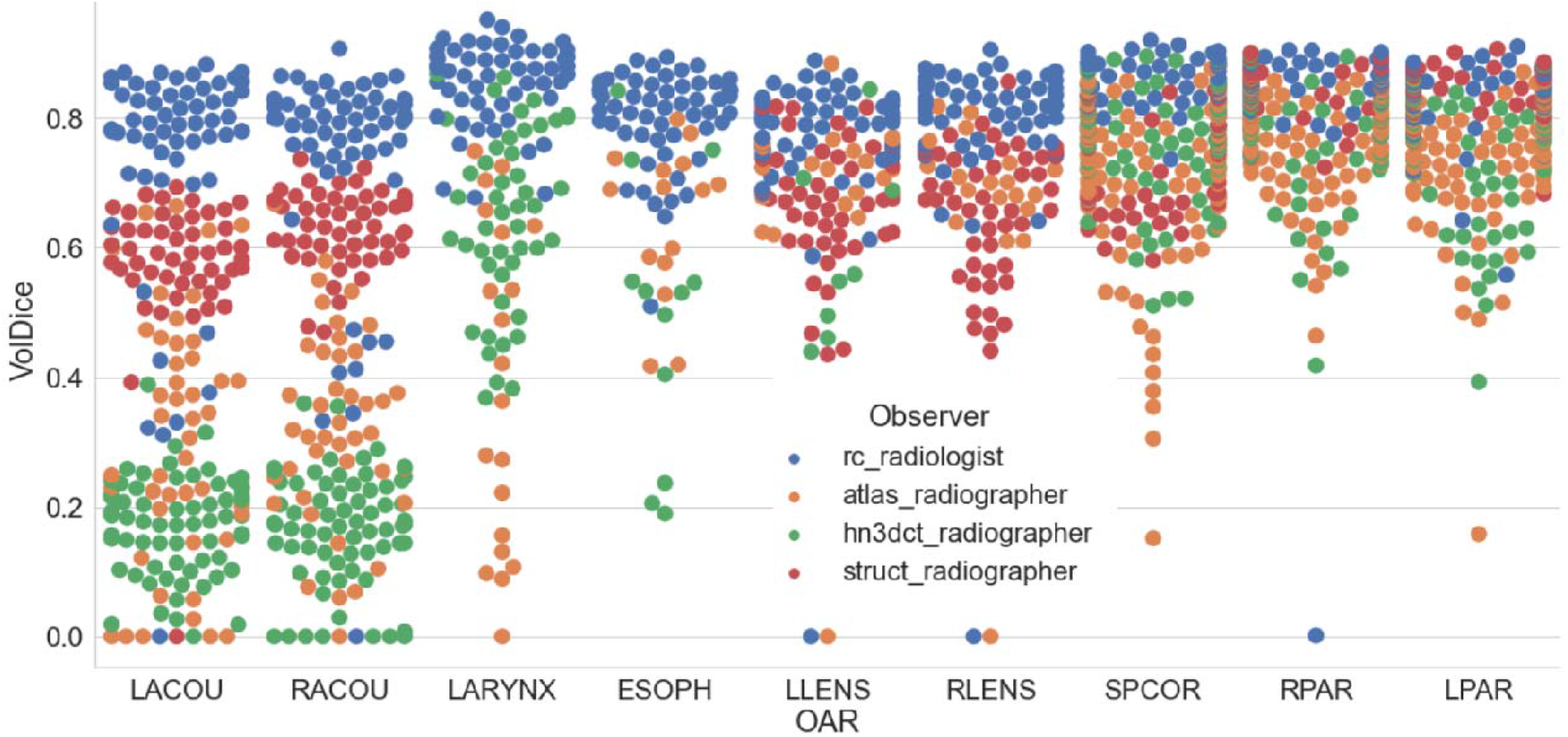
Performance differences of WOLNET ensemble for select OARs on external datasets. Beeplot to show variation of 3D Volumetric DICE performance of WOLNET ensemble for select OARs (L/R Acoustics, Larynx, Oesophagus, L/R Lens, Spinal Cord, L/R Parotids) across different datasets. Broad spectrum in contouring protocols of the acoustics across different centers.

### Generalizability Assessment

To assess the generalizability of the WOLNET model, we used 7 external datasets that have been generated in different institutions (Figure 1, Supplementary Table 1). While external datasets are valuable to assess generalizability of AI methods, these datasets released by external centres had variable labels of OARs that overlapped with our analysis (Supplementary Table 2). In addition to this, the quality of the “Ground-Truth’’ labels for each dataset are only as good as the observer generating the labels, and therefore it is important to note that there may exist variability within contouring protocols for multiple OARs generated at these independent centres. We found extensive variability of ground truth information extracted from each external dataset and only a subset of OARs segmented in this study overlapped with any given dataset (Supplementary Table 2). One dataset Radiomics-HN1 (RHN1) had the most overlapping OAR categories with our RADCURE dataset (17 out of 19 OARs successfully overlapped). TCIA-HNSCC (TCHN) had the least overlapping categories (5 out of 19 OARs). Results for each external dataset can be found in Appendix B (Supplementary Figure 8, Supplementary Tables 4 and 5) [5,20,43–52].

## Discussion

In this study, we introduce SCARF, a six-step benchmarking framework for evaluating open-source AI models’ performance and clinical acceptability in auto-segmentation of essential radiation therapy targets for head and neck cancer treatment. Our results show the majority of OARs generated by the best model tested required only minor edits for use in radiation therapy plans. SCARF proves to be a valuable framework for open-source resources’ curation, model training, performance evaluation, and clinical acceptability testing, providing a protocol for recording baseline results for each OAR category’s segmentation performance.

SCARF, with its focus on easy and reproducible benchmarking of auto-segmentation systems, can significantly improve the AI evidence pyramid in the context of radiation therapy planning. By providing open access to all data and methods used in the analysis, SCARF addresses some of the key challenges faced in the field of radiation therapy and AI integration. In the context of the AI evidence pyramid [16], SCARF can play a vital role in the external validation of AI models for auto-segmentation in radiation therapy as shown in the context of OAR segmentation for HNC. The availability of preprocessed datasets and open access to all relevant information ensures that AI models can be rigorously evaluated using different patient populations and clinical scenarios. This process enhances the reliability and generalizability of AI models, moving them closer to the third step of the AI evidence pyramid.

Moreover, the transparency provided by SCARF enables better assessment and benchmarking of segmentation systems, which is crucial in moving towards the creation of usable AI tools (the fourth step of the pyramid) in radiation therapy planning. Researchers and clinicians can examine the methods and results of the study, enabling them to build upon the findings and implement the technology in clinical practice more confidently. Furthermore, SCARF’s emphasis on clinical acceptability testing aligns with the notion of optimising networks not only for model performance but also for their practicality in a clinical environment. This focus is in line with the goal of increasing the number of randomised clinical trials (the penultimate step of the pyramid) to evaluate the true impact of AI-driven auto-segmentation systems in real-world radiation therapy scenarios. By ensuring that AI models are not only accurate but also meet the requirements of clinical experts, SCARF facilitates the integration of AI technology into routine clinical workflows and can be used as an introductory code base to facilitate compliance with the checklists governing proposing AI interventions for auto-segmentation in radiation therapy of head and neck cancer (Table 2) providing an allotted time savings of over 450 developer hours.

**Table 2:**
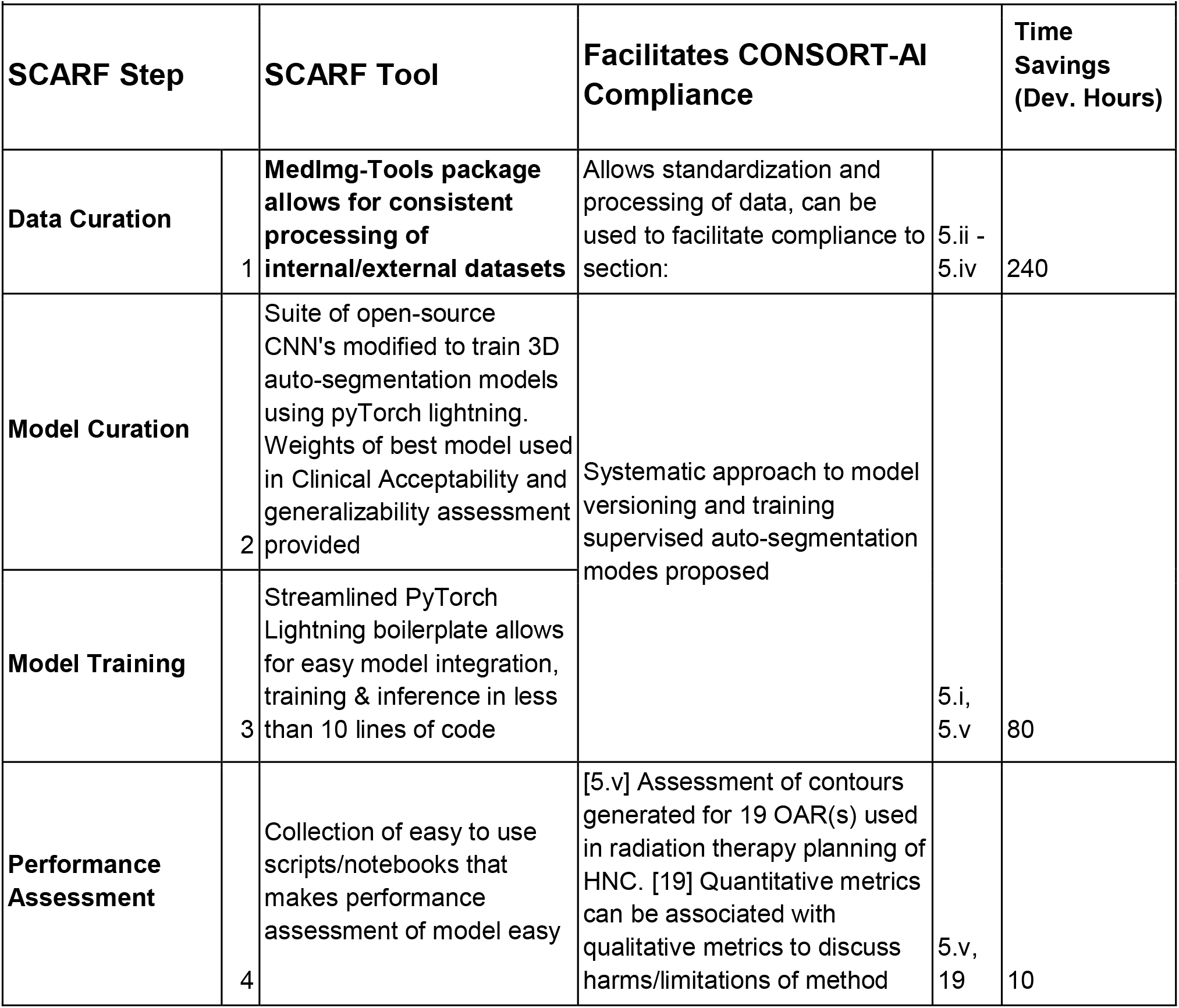
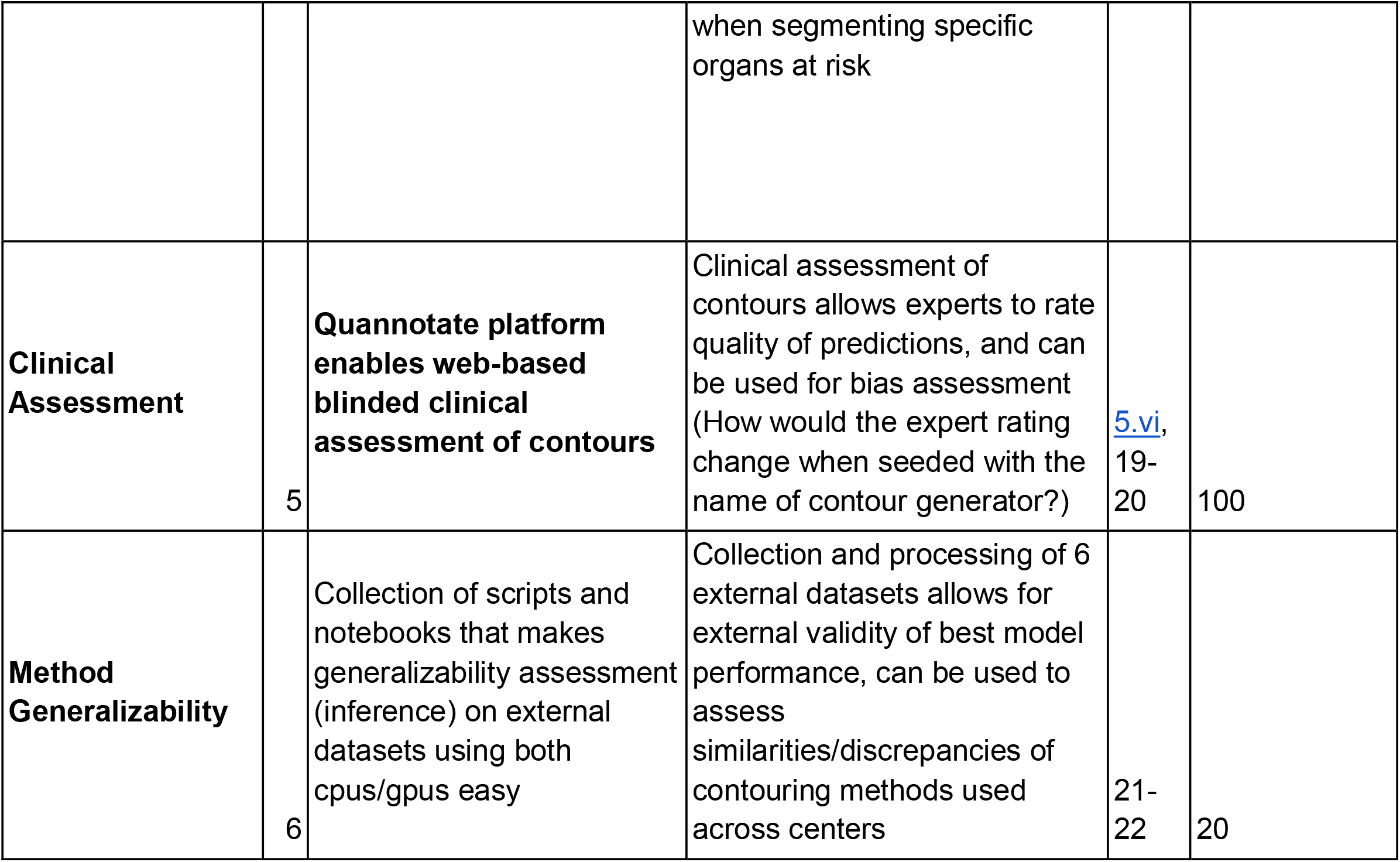
SCARF’s toolkit can be used to facilitate compliance to checklists like that provided by CONSORT-AI [15].

This study has several limitations. The open-source repositories used in the analysis were collected and trained up to April 2020, missing potential newer models and opportunities for further analysis. The primary focus was on proposing a reproducible training framework for auto-segmentation pipelines, not on quantitative superiority. The clinical assessment step involved only four radiation oncologists due to resource constraints, limiting statistical significance. Acceptability ratings showed significant differences in only three out of thirteen organ-at-risk categories, suggesting the need for further analysis with more clinicians. The Quannotate clinical testing interface provided valuable insights but may benefit from exploring other assessment methods. The models trained were restricted to datasets with complete ground truth labels, limiting their usability with datasets containing partial labels. Future work should address this limitation and explore improvement opportunities using SCARF as a benchmark for effectiveness.

## Conclusion

In conclusion, SCARF, our open-source and reproducibility framework for auto-segmentation in radiation therapy planning, significantly enhances the AI evidence pyramid in this medical domain. By promoting transparency, facilitating external validation, and emphasising clinical acceptability, SCARF enables robust validation studies by multidisciplinary teams, bridging the gap between AI research and clinical practice. This advancement can lead to improved standards of care for radiation therapy patients and open up new avenues for research and advancements in the field. Our study also highlights the importance of incorporating both quantitative and qualitative controls in benchmarking auto-segmentation systems. With SCARF, we provide a comprehensive six-stage framework that enables benchmarking state-of-the-art convolutional neural networks against essential organs-at-risk in head and neck cancer. The availability of SCARF and the clinical assessment toolkit fosters transparency, reproducibility, and acceptance of auto-segmentation systems in clinical practice, accelerating the adoption of reliable and efficient models in radiation therapy planning and beyond.

## Notes

### Competing Interest Statement

Benjamin Haibe-Kains is a shareholder and paid consultant for Code Ocean Inc. There are no conflicts of interest.

### Clinical Protocols

https://www.quannotate.com

### Funding Statement

This study was funded by Canadian Institutes of Health Research Project-Scheme for the Development and comparison of radiomics models for prognosis and monitoring (Haibe-Kains B, Hope A). Term: 02/2020-01/2023

### Author Declarations

A UHN institutional review board approved our study and waived the requirement for informed consent (REB 17-5871); we performed all experiments in accordance with relevant guidelines and ethical regulations of Princess Margaret Cancer Center. Only PM data used for network training was involved in the REB. External publicly available datasets were governed by individual REBs by its institution of origin.

### Summary of Updates

Updates were made to manuscript re-fotmatting for the proposed framework. Changes are reflected in all sections. Abstract and Discussion have been updated to reflect changes. Figure 1 was moved to suplementary material, and replaced by an overview of the proposed framework. Figure 2 was changed to emphasize Model Curation and Model training steps in the reproducibility stage. Figure 3 was updated by adding components of Supplementary Figure 2 to make a clearer representation of the 5th step in the framework, clinical assessment. Figure 4 was changed entirely and shows performance of trained ensemble for select OARs as a representation of the 6th step (Generalizability Assessment). Supplementary Table 4 was moved to Table 1. Table 2 was added to show potential impact of SCARF when used to cohere to pre-published AI checklists. A 7th external dataset was added to generalizability assessment.

