## Supplementary material for "SCARF: Auto-Segmentation Clinical Acceptability & Reproducibility Framework for Benchmarking Essential Radiation Therapy Targets in Head and Neck Cancer": Suplemetary Materials

**Appendix**

Manual delineation of radiation therapy targets is an essential, yet time consuming process. In radiation therapy, three major categories or regions of interest are usually delineated before dose assignment and planning optimization. These include gross tumour volumes (GTVs), clinical target volumes (CTVs) where radiation dose is prescribed and normal organs noted organs-at-risk (OARs). Accurate OAR contouring by radiation technicians on planning CT scans is heightened when treating head-and-neck cancers (HNC) where excess radiation dose delivery during therapy could propel the occurrence of radiation induced toxicity after treatment including xerostomia (reduced salivary output affecting salivary and parotid glands) and dysphagia (complications affecting facial and neck muscles involved with swallowing) [[1]](https://paperpile.com/c/v2Np13/mEycO). Patients with these conditions have lower quality of life and are at an increased risk of suicide [[2,3]](https://paperpile.com/c/v2Np13/e0oXz+XM81w). In order to most actively ensure that radiation induced toxicities and negative effects to patient quality of life are minimised, accurate and complete delineation of organs in the treatment field is necessary while maintaining maximum tumour control. The difficulty of this task cannot be understated due to the size, volume and close proximity of the OARs being contoured in the head and neck region. In total, there are over 40 OARs that are recommended to be delineated by current consensus guidelines [[4]](https://paperpile.com/c/v2Np13/FFYT1). Consequently, it can take up to 180 minutes for the clinical team to contour all OARs required in any given treatment plan [[5]](https://paperpile.com/c/v2Np13/CLU3H).

Early deformable atlas-based auto-segmentation approaches [[6–11]](https://paperpile.com/c/v2Np13/I8lha+UuYky+NCHAt+8AK2R+4xgGu+E1RcS) have been superseded by more precise deep learning-based approaches using convolutional neural networks (CNN). Variations of architectures have held the state-of-the-art for automated segmentation of OAR contours in the head and neck region since 2016 [[12–24]](https://paperpile.com/c/v2Np13/zdkiC+jofLO+HjoMS+LWAT9+qGAYp+8jkNo+NWZBe+WMgGQ+g89cX+HkkIy+ZNnkc+eGoAl+A23Ka). Recent publications have shown the utility of properly trained and optimised simple CNN architectures when applied to complex auto-segmentation tasks [[25]](https://paperpile.com/c/v2Np13/YWPDQ). This suggests that model complexity does not necessarily correlate with increased model performance, while method configuration affects performance at a higher level than structural architectural variations.

### **Appendix A: Methods & Materials**

#### **Dataset Curation**

Our data processing revealed a total of 34 OARs in the head and neck region that had been contoured over the study period. Naming of targets in RT structure files were standardised and extracted in a reproducible manner using a documented script (see Code Availability section). Patients were selected for this analysis if they had all of a selected amount of OAR(s) deemed essential by expert radiation oncologists delineated in their RT-STRUCT DICOM file. The resulting dataset is referred to as RADCURE [[26]](https://paperpile.com/c/v2Np13/maymw). To complement our RADCURE internal medical imaging datasets, we curated a large compendium of external datasets made publicly available via TCIA. Similarly to RADCURE, we used our Med-ImageTools package to curate and harmonise this data compendium to test the generalizability assessment of the auto-segmentation models investigated in this study [[27]](https://paperpile.com/c/v2Np13/ZOEFe). External publicly available datasets were governed by individual REBs by their institution of origin.

#### **Exclusion Criteria For Model Selection**

Studies were excluded from this analysis based on code availability, computational capacity, model integrability and repository maintenance. More specifically, we excluded 2D architectures whose convolutional schemes could not be directly updated to 3D without disrupting architectural integrity. Primary selection criteria included models originally written in PyTorch [[28]](https://paperpile.com/c/v2Np13/Qk0a6), that were 3D or could be minimally modified to accept 3D inputs. Popularity of repositories on GitHub quantified by its public GitHub star rating was also a factor used when selecting architectures with similar architectural layouts. Selected networks were to be trained end-to-end directly, with no complex architectural or computational restrictions and with minimal modifications made to original source code provided by the original authors to keep architectural integrity.

#### **Model Training**

We combined a modified and weighted TopK Cross entropy loss [[16]](https://paperpile.com/c/v2Np13/qGAYp), which selects the top 10% of most difficult voxels (defined by the largest loss values) in the weighted cross-entropy loss calculation and only added the contribution of these voxels to the loss with the Focal Tversky Loss, which has been previously defined to excel at semantic segmentation tasks where pixel-wise class imbalance between classes is high [[29,30]](https://paperpile.com/c/v2Np13/kB7Pm+s9JQG). The RADAM optimizer was used to minimise the loss during training [[31,32]](https://paperpile.com/c/v2Np13/cXPAf+4oTcT). The initial learning rate of the optimizer was set to .001. The tuning loss was monitored and the learning rate decreased by a factor of 0.5 if there was no significant change in tuning loss after 12 epochs. The voxel spacing of each patient scan was standardised to a lower resolution of 1mm x 1mm x 3mm using SimpleITK [[33]](https://paperpile.com/c/v2Np13/jRvKu).

An augmentation scheme was implemented during training. Translation, mirroring, zooming, and rotation were applied in-plane (x-y plane only) and at random during training. We used random translations between -32 and 32 pixels in each plane; uniform scaling factors between 0.9 and 1.1; and mirrored images with a probability of 0.5. Patient scans were cropped to a size of (192x192x64) pixels for use during training. The ground truth mask was first used to identify the coordinates of the patient’s centre of mass on which cropping was based. All CT volumes used in analysis were clipped to a HU range of -200 to 300 (a common soft tissue window applied by radiation oncologists) before augmentations were applied. Z-score normalisation was applied after all augmentations were completed.

#### **Model Fine-tuning**

To boost performance, we selected the best model for a 2nd re-training phase on full-resolution patient scans for an additional three days on 4 NVIDIA Tesla V100 GPUs and tested using MONAI based sliding-window inference [[34]](https://paperpile.com/c/v2Np13/1gKZq). We set the initial learning rate to 4e-4, tuning loss was monitored and the learning rate decayed by a factor of 0.96 after no change in loss for 1 epoch while all CT volumes used in the final analysis were clipped to a HU range of -500 to 1000. Cropping regimen was modified to the size of (192x192x128) pixels. Images were not resampled for re-training. We used random translations between -64 and 64 pixels while the other randomised augmentations and Z-score normalisation scheme were kept unchanged. The initial convolutional setting of the best network was set to 48 feature maps instead of 32 feature maps used during the tuning phase of our study. Although large parameter dense CNNs could lead to overfitting and subsequent poor generalizability when data used for training is sparse [[35]](https://paperpile.com/c/v2Np13/dKrEz), Performance improvements can be obtained by adding feature maps at each network layer. This increases the model’s capacity to learn complex relationships like learning to segment complex anatomical features from a medical image.

Ensembling is one method to boost model robustness and has been frequently employed across a wide range of medical image-based auto-segmentation tasks [[36–40]](https://paperpile.com/c/v2Np13/sKNq8+pSeoX+1YUHu+Ly8tc+YkZfa). To minimise variance and spurious model predictions, we used five random K-Fold training/tuning splits to build a “Fine-tuned” version of WOLNET to increase the generalizability potential of the final model [[41]](https://paperpile.com/c/v2Np13/rwApz). The training scheme was modified as follows: random crops of size (112px x 176px x 176px) were used to train the network with a batch size of 2. Each fold was trained on 4 NVIDIA Tesla V100 GPUs with 32G RAM, on the original multi-class segmentation task for 3 days or until convergence. Since there is heterogeneity in the patient population used during training, scans extracted have variable amounts of slices.

#### **Feature Description Of Quannotate Update**

Observers were able to access the contours online from different locations and were blinded to the assessments by other observers. In this study, we re-engineered QUANNOTATE, an open-source cloud deployed quality assurance tool developed to conduct blinded assessment of radiation therapy contours by expert radiation oncologists [[42]](https://paperpile.com/c/v2Np13/D7sMV). The web application was modified so that each user is able to assess specific contours by individual OAR category. A contour evaluation protocol was defined and implemented to determine clinical validity of the contours produced by the best method when quizzing experts. Inside the test dashboard, observers are able to slide through an entire patient scan, in an OAR dependent manner, using the integrated in-browser window slider. Users can select between different clinical CT windowing levels of the patient’s CT image where the contours have been superimposed. There are currently three windowing level settings that can be used during the assessment, these include bone, soft tissue and lung views.

Users were asked a series of questions about each OAR in the contour set. The first question, “Who delineated this contour?” with options “I don’t know”, “Human” or “Computer”. The next question assesses the clinical acceptability by getting the user to ‘rate’ the contour being assessed. We adopted an established 5 point rating clinical acceptability scale for our study [18,48]. The contour acceptability scale ranged from 1 (Poor, large areas need minor or major edits, is unusable for planning purposes), 2 (Fair, needs significant edits to be used for planning purposes), 3 (Good, needs minor edits to be used for planning purposes), 4 (Within acceptable inter physician variation for planning purposes) and 5 (Perfect, indistinguishable from physician drawn contours for planning purposes). The users were blinded to the origin of the contour at the time of the questionnaire, and answers were exported to be analysed after all users completely assessed all contours. Individual ratings of 4 or higher will be considered acceptable, the condition being no additional edits are required to transition set contour into a radiation therapy plan. When analysing ratings averaged across all observers, a contour will be considered clinically acceptable when mean acceptability rating (MAR) for that contour across all observers is above 3.5, and not-clinically acceptable when MAR is less than 3.5. (Supplementary Figure 2)

#### **Model For Generalizability Assessment**

We simulate a “clinic-like” environment for external validation by using no ground-truth information to identify the centre of mass of any scan during inference. During inference, Otsu’s thresholding [[43]](https://paperpile.com/c/v2Np13/t9g1l) was used to first create a ‘mask’ of the body, from which the patient’s centre of mass was calculated and used to crop the image in the x-y plane with dimensions of 292x292px (Original dimensions of a Radcure scan in the x-y plane are 512x512, the relevant anatomy is found in more than half of the image, hence image cropping boosts inference efficiency). Sliding window inference was applied to the cropped version of the patient scan. Final predictions were averaged across models trained from/on each of the five folds and performance on each external dataset was assessed using the same segmentation metrics as in the training phase.

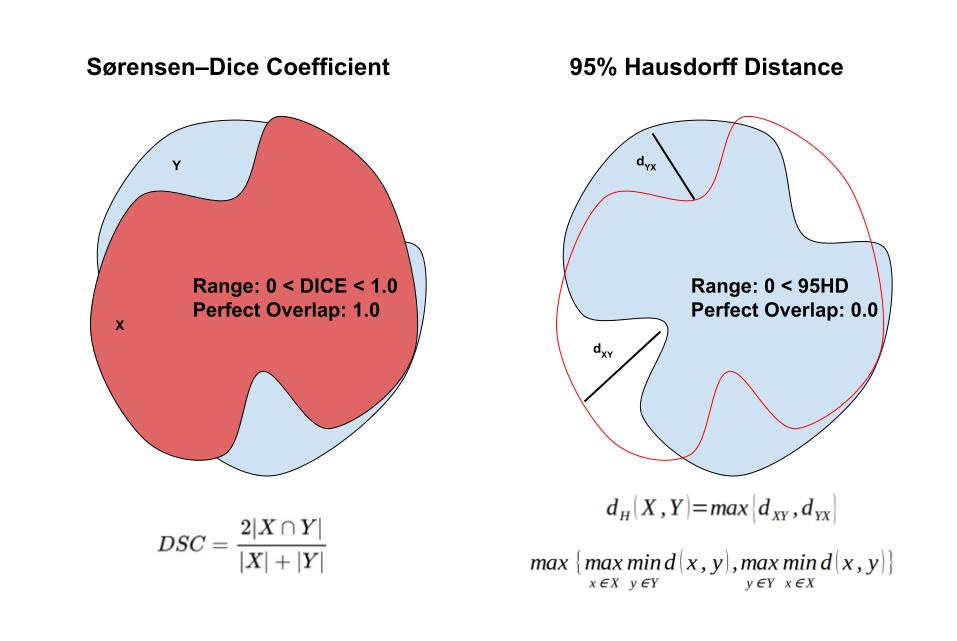

###### **Supplementary Figure 1: Performance Based Overlap Metrics**

(Right) Shows a 2D example of Sorensen-Dice coefficient Calculation (DICE), in this study we use a volumetric dice overlap metric where Y is the sets of foreground voxels in the ground truth annotations and X is the set of foreground voxels produces by the deep learning auto-segmentation network. (Left) Shows a 2D example of Hausdorff distance, our calculation is based on the 95th percentile of the distances between the boundaries of volume X and Y, this minimises the impact a small subset of outliers could have on distance calculation.

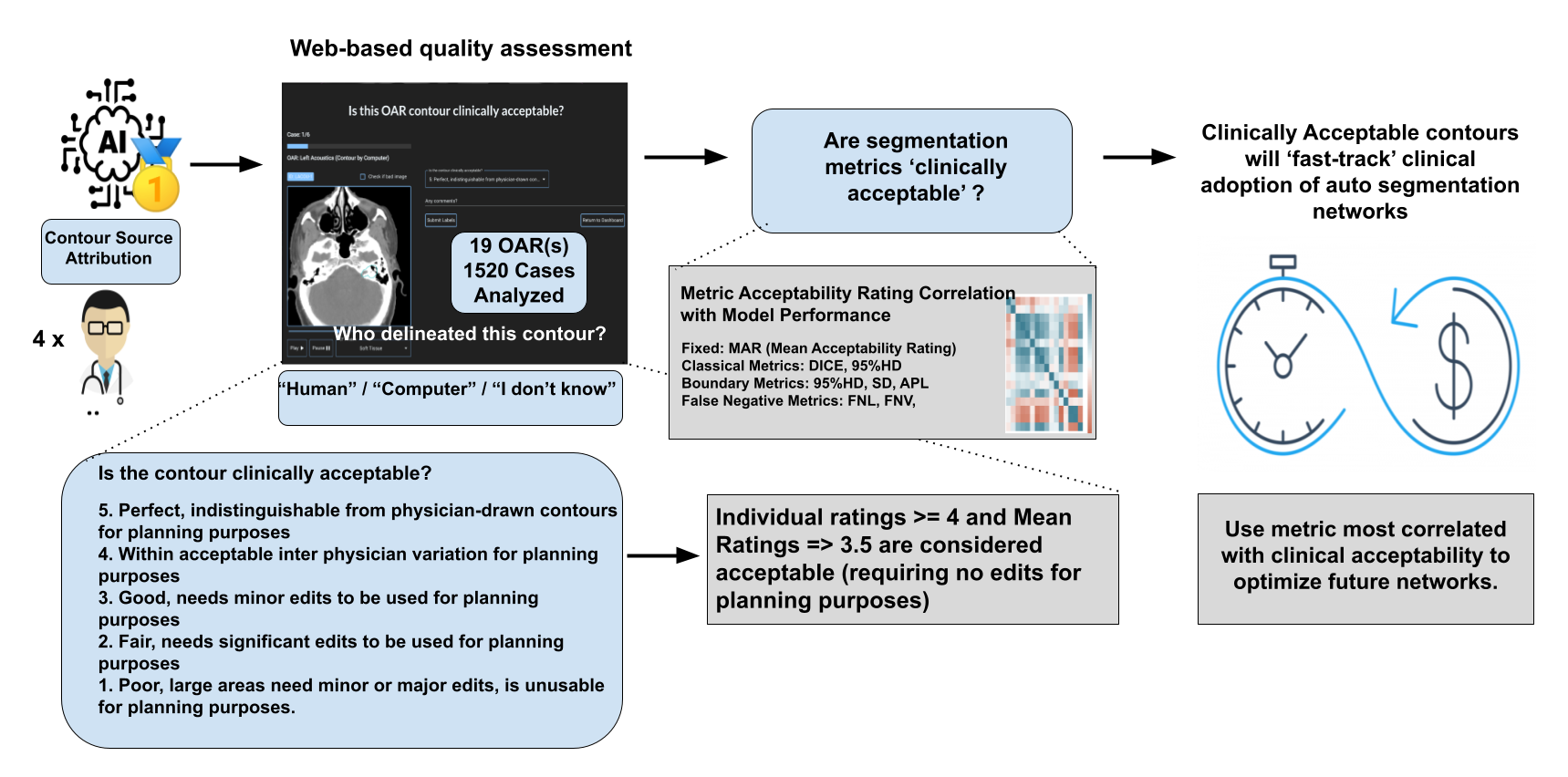

###### **Supplementary Figure 2: Overview of Clinical Acceptability Protocol, an open source web-based quality assurance tool from our previous study was modified for clinical acceptability testing of radiation therapy contours.**

For this analysis, 4 expert radiation oncologists were each given the opportunity to assess the acceptability of deep learning or manual ground truth contours in a blinded fashion. 10 ground truth contours paired with 10 deep learning generated contours for the same patient were extracted for each of the 19 OARs. A total of 380 3D contours were assessed by each observer. They were asked to rate acceptability on a 5 point scale taking the complete volume of the entire OAR contour into context. A rating of 4.0 and higher can be considered ‘acceptable’ in that no edits are required by the examining physician for planning purposes. They were also asked to ‘guess’ the observer that generated the contour (“Human”/ “Computer” / “I don’t Know”) before submitting their rating. Mean Acceptability Ratings were then calculated for each OAR, and analysis assessing correlation of acceptability with 6 different segmentation performance metrics was conducted. These metrics include 3D Volumetric Dice Overlap Coefficient, 95% Hausdorff Distance, Applied Path Length, False Negative Length, and False Negative Volume

### **Appendix B: Results**

#### **Contents of Radcure Cohort Used In Analysis**

The study cohort consisted of 378 oropharyngeal, 123 nasopharyngeal, 10 hypopharyngeal, 10 oral cavity, and 7 laryngeal cancer patients as well as 55 patients of unknown or other primary-site cancers. The 19 OARs included acoustics (L/R), brachial plexuses (L/R), brainstem, chiasm, oesophagus, eyeballs (L/R), larynx, lenses (L/R), lips, mandible, optic nerves (L/R), parotid glands (L/R), and spinal cord. This subset of RADCURE was randomly divided once into training, tuning and testing divisions. 10% of this cohort (59 scans) were selected at random and set aside to be used as an independent testing set. Out of the remaining 523 patients, 44 patients were used for tuning and 479 patients were used for training each segmentation network (Supplementary Figure 4).

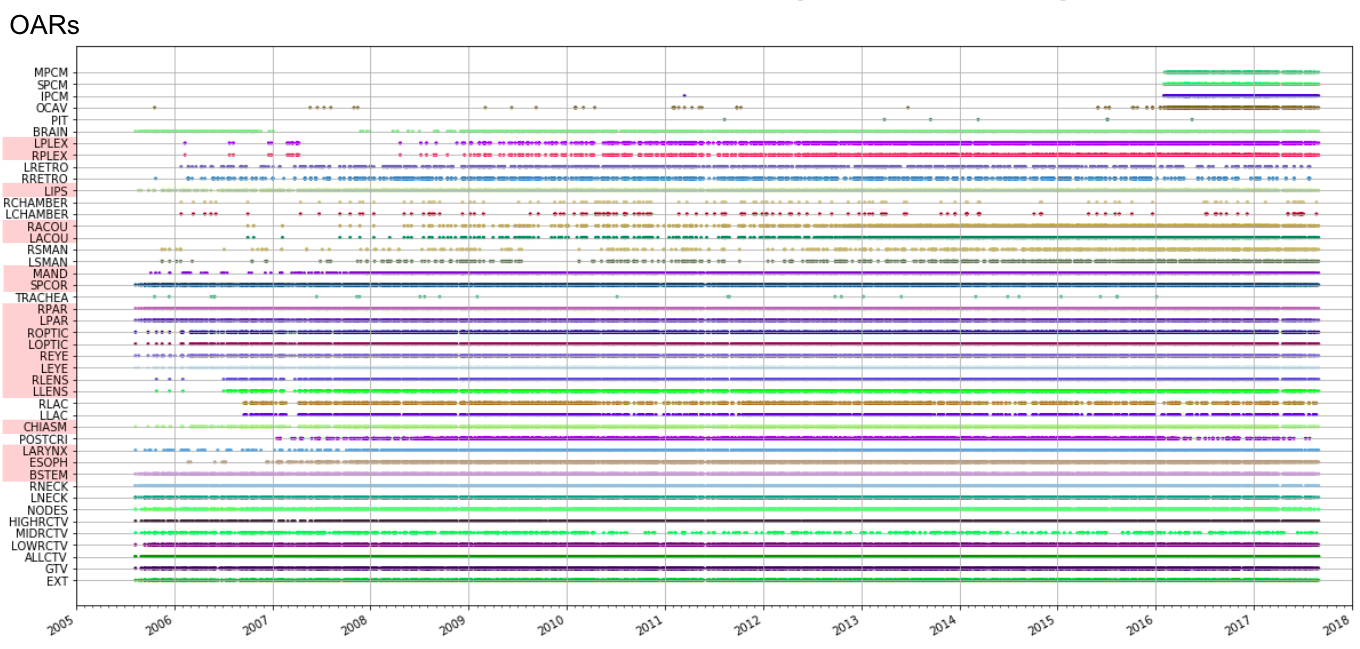

**Supplementary Figure 3:** Availability of OAR segmentation in our cohort of 3,3489 HNC patients included in our RADCURE dataset. A subset of 582 patients have 19 OARs fully segmented (highlighted in red), this set was used to train and validate our compendium of 11 open-source auto-segmentation models (see preliminary results).

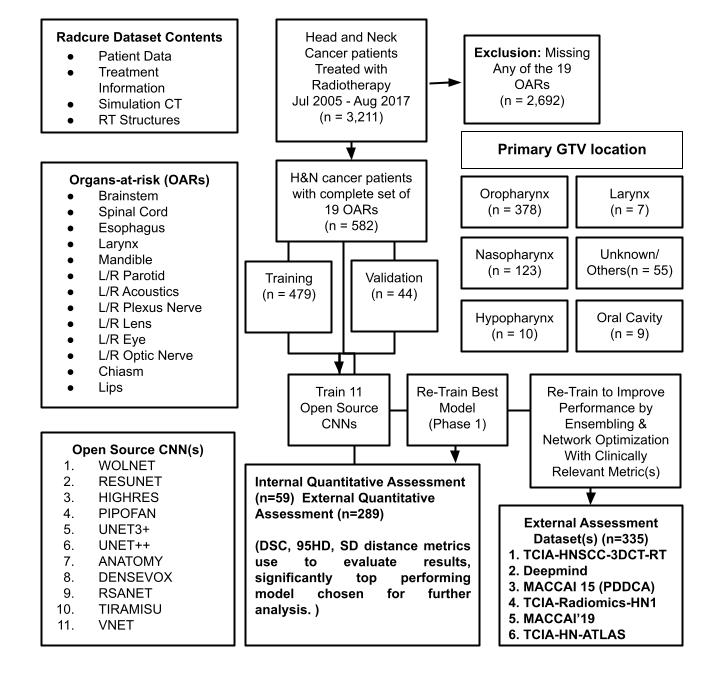

###### **Supplementary Figure 4: Cohort Diagram depicting patient & OAR selection for our study.**

A total of 582 patients were selected for this analysis because they had all of 19 OAR(s) deemed essential by expert radiation oncologists delineated in their RT structure dicom file. A series of 11 open-source architectures were selected for use and trained to segment each of the OARs listed simultaneously. Performance metrics (DICE, 95HD) allowed us to select the best model to re-train for clinical acceptability analysis which was conducted on the QUANNOTATE web platform. Acceptability testing allowed us to draw correlations between quantitative and qualitative results. For external validation, an ensemble of the best network would be trained with a modified loss which adds more weight for performance metrics most closely correlated to clinical acceptability. This model ensemble would then be applied to 6 datasets collected for external validation.

| **Supplementary Table 1. External HNC imaging and segmentation datasets.**  Missing data are represented in gray. ‘Available Data’ types that are only available for a portion of patients are represented by a transparently coloured cell. | | | | | | | |
| --- | --- | --- | --- | --- | --- | --- | --- |
| Dataset | Institution | Scans | Available Data | | | | References |
|  |  |  | Imaging | Clinical | OAR | GTV |  |
| Radcure | UHN | 2552 |  |  |  |  | [[44,45]](https://paperpile.com/c/v2Np13/GR4gX+Ql1G) |
| HNSCC-3DCT-RT | MIAMI | 94 |  |  |  |  | [[46,47]](https://paperpile.com/c/v2Np13/3OS3A+OhTf) |
| Deepmind | HMS+Multi-Site | 35 |  |  |  |  | [[16]](https://paperpile.com/c/v2Np13/qGAYp) |
| PDDCA | HMS | 48 |  |  |  |  | [[48]](https://paperpile.com/c/v2Np13/PXGkO) |
| Radiomics-HN1 | MAASTRO | 137 |  |  |  |  | [[49–51]](https://paperpile.com/c/v2Np13/6AkMC+AJzS+bP62) |
| STRUCTSEG19 | CAS | 50 |  |  |  |  | [[52]](https://paperpile.com/c/v2Np13/Bzp7J) |
| Head-Neck-CT-Atlas | MDACC | 215 |  |  |  |  | [[53,54]](https://paperpile.com/c/v2Np13/XLeAe+o3uY) |
| SegRap 2023 | UESTC | 120 |  |  |  |  | [[55]](https://paperpile.com/c/v2Np13/Jb1H) |

| **Supplementary Table 2: Distribution of Ground Truth OAR labels (masks) in external datasets** | | | | | | | | | |
| --- | --- | --- | --- | --- | --- | --- | --- | --- | --- |
| **OAR** | **RADCURE (n=59)** | **HNSCC-3DCT-RT (n=94)** | **Deepmind - Onc (n=35)** | **Deepmind- Rad (n=35)** | **PDDCA (n=48)** | **TCIA-HNSCC (n=215)** | **STRUCTSEG(n=50)** | **Radiomics-HN1 (n=137)** | **SegRap2023 (n=120)** |
| **BSTEM** | 59 | 82 | 35 | 35 | 48 | 0 | 50 | 128 | 120 |
| **CHIASM** | 59 | 61 | 0 | 0 | 48 | 0 | 50 | 13 | 120 |
| **ESOPH** | 59 | 12 | 0 | 0 | 0 | 0 | 0 | 15 | 120 |
| **LACOU** | 59 | 0 | 0 | 0 | 0 | 0 | 50 | 46 | 120 |
| **LARYNX** | 59 | 47 | 0 | 0 | 0 | 0 | 0 | 20 | 120 |
| **LEYE** | 59 | 46 | 35 | 35 | 0 | 0 | 50 | 11 | 120 |
| **LIPS** | 59 | 5 | 0 | 0 | 0 | 0 | 0 | 1 | 0 |
| **LLENS** | 59 | 9 | 35 | 35 | 0 | 0 | 50 | 21 | 120 |
| **LOPTIC** | 59 | 27 | 35 | 35 | 48 | 0 | 50 | 11 | 120 |
| **LPAR** | 59 | 71 | 35 | 35 | 48 | 94 | 50 | 112 | 120 |
| **LPLEX** | 59 | 0 | 0 | 0 | 0 | 0 | 0 | 0 | 0 |
| **MAND** | 59 | 60 | 35 | 35 | 48 | 0 | 50 | 74 | 120 |
| **RACOU** | 59 | 0 | 0 | 0 | 0 | 0 | 50 | 42 | 120 |
| **REYE** | 59 | 49 | 35 | 35 | 0 | 0 | 50 | 10 | 120 |
| **RLENS** | 59 | 0 | 35 | 35 | 0 | 0 | 50 | 20 | 120 |
| **ROPTIC** | 59 | 28 | 35 | 35 | 48 | 0 | 50 | 10 | 120 |
| **RPAR** | 59 | 71 | 35 | 35 | 48 | 93 | 50 | 116 | 120 |
| **RPLEX** | 59 | 0 | 0 | 0 | 0 | 0 | 0 | 0 | 0 |
| **SPCOR** | 59 | 83 | 35 | 35 | 0 | 119 | 50 | 132 | 120 |

#### Model Curation

Many networks cannot be reproduced easily because of various computational or organisational hurdles. In addition, many publications fail to compare findings to validated baseline models. Complex network modifications may only prove useful if they are compared to a simple baseline architecture. To address the lack of open source resources to validate findings of previously published studies, we performed a large comparative analysis of open-source deep neural networks applied to image segmentation in a medical context. We looked for additional studies published on a medical image segmentation task to collect open-source networks that could be trained on our OAR segmentation task. A total of 2D networks that could not be converted to 3D convolutional scheme without modifying the architectural integrity, close similarity or overlap between other architectures, code released with the study was unmaintained and/or could not be integrated into PytorchLightning.

| **Supplementary Table 3: Architectures selected for integration to SCARF code-base for easy re-implementation, training & validation.** | |
| --- | --- |
| **Name** | **Description** |
| **3D-UNET (WOLNET)** | One popular pytorch implementations of the standard 3D UNET paper [[56]](https://paperpile.com/c/v2Np13/0RXHK) were taken and used as the baseline architecture. Named WOLNET after it’s author. [[57,58]](https://paperpile.com/c/v2Np13/38JFz+fmGCe) |
| **3D-RESUNET** | We integrated a third party implementation [[57,58]](https://paperpile.com/c/v2Np13/38JFz+fmGCe) of a residual symmetric 3D UNET proposed by [[59]](https://paperpile.com/c/v2Np13/q88Oc). They introduced a residual skip connection to each valid convolutional module present in their network. To minimise information loss, this network does not down sample feature maps along the z dimension. To minimise the effects of anisotropy 2D convolutions are used in the modules at the lowest part of the network, which contain fine scale feature maps. Each regular residual module will apply in total 7x7x5 of nonlinear convolutions to the input. (To embed 2D features 3x3x1 convolutions are applied followed by subsequent 3x3x3 convolutions.). |
| **HIGHRESNET** | Li, W et al (2017) choose to integrate dilated convolutions and residual connections in their proposed 20 layer residual network. These residually connected dilated convolutions allowed for multi-scale feature preservation as training progressed. [[60]](https://paperpile.com/c/v2Np13/QFAZb) A third party implementation of HighResNet3D was used for our analysis. [[61]](https://paperpile.com/c/v2Np13/XeUfu) |
| **PIPOFAN** | We modified the original 2D Pyramid Input Pyramid Output Network proposed by Fang, et al (2020) to accept 3D volumes. This 3D pyramid abstraction network (PIPOFAN) processes the volumetric input by applying 3D Equal Depth Convolutions (EDC), after passing through the network the pyramid outputs are fused together which has been shown to improve subsequent organ segmentations. This network was originally used to segment multiple thoracic OARs on individual slices of a CT scan. [[62,63]](https://paperpile.com/c/v2Np13/e0Uxj+tws2b) |
| **UNET3+** | We introduced a 3D version of a 2D UNET3+ proposed by Huang, H et al (2020) which was created as a modification of the UNET++ that incorporated full-scale skip-connections into the UNET++ network. This architecture was engineered to produce full-scale aggregated feature maps that allows deep supervision components to learn more comprehensive hierarchical feature maps with the hopes of producing more accurate contours. [[64,65]](https://paperpile.com/c/v2Np13/xrGf6+ZalTn) |
| **UNET++** | We integrated a 3D version of the 2D Nested Unet Architecture (UNET++) proposed by Zhou et al (2018) using their code as the base network for the architecture used in the study. The authors redesigned sip connection pathways of the original UNET architecture with the intent to reduce the semantic gaps between the encoding and decoding feature maps. This is the first paper to propose and integrate deep supervision into their network where the outputs of each individual segmentation branch are averaged before the final softmax layer of the network. [[66,67]](https://paperpile.com/c/v2Np13/s9ddl+DNGF3) |
| **ANATOMY** | AnatomyNet was the only segmentation model used in this analysis that was previously published on a HNC OAR segmentation task. This UNET variant incorporates squeeze and excitation residual building blocks in the downsampling/upsampling layers of the network. Code was refactored and updated to suit the newest versions of pytorch. [[18,68]](https://paperpile.com/c/v2Np13/wDRjn+NWZBe) |
| **DENSEVOX** | A pytorch based third party implementation of DenseVoxNet 3D first proposed by Yu, L et al 2017, for cardiac segmentation was integrated into our study. This network consists of two DenseBlocks in the downsampling part of the network which are densely connected. In total there are 24 transformation layers before upsampling. A long skip connection was used to stabilise the training process by connecting the transition layer to the output layer. [[69,70]](https://paperpile.com/c/v2Np13/vzQuz+JyDjn) |
| **TIRAMISU** | We adapted a third party 2D implementation of a fully convolutional Densenet originally presented by Jegou et al, 2017 (named 100-layer tiramisu). This paper was the first to apply the DenseNet to the problem of 2D semantic segmentation. Densenets are constructed by concatenating each output of a subsequent densely connected convolutional block to the next block, therefore linearly augmenting the number of feature maps after each ‘down transition’. This does not occur in the upsampling part of the network. The feature maps from the downsampling path are then concatenated with those of the upsampling path to produce a predicted segmentation mask at the resolution of the original input. [[71,72]](https://paperpile.com/c/v2Np13/SbwSf+SA3QS) |
| **RSANET** | RSANet is a 3D recurrent slice-wise attention network proposed by Zhang, H et al (2019) could be directly integrated into our network. Originally constructed for Multiple Sclerosis lesion segmentation this network utilises slice wise attention blocks to help capture long-range inter-slice dependencies along any direction of a 3D medical image. These blocks allow for the recurrent aggregation of information along multiple directions therefore providing a mechanism to help capture global contextual information, which can be used to produce more accurate segmentations. [[73,74]](https://paperpile.com/c/v2Np13/xIOzF+ROLib) |
| **VNET** | An updated third party implementation of 3D VNET architecture proposed by Millerari et al (2016) was used in this study [[75]](https://paperpile.com/c/v2Np13/UfgJw). The VNET architecture is a fully convolutional network based on the original UNET. The authors choose to replace 3x3x3 convolutions present in UNET by 1 strided 5x5x5 convolutions. Additionally, in place of max-pooling, 2x2x2 convolutions with stride of 2 were used during down sampling. Finally, PReLU nonlinearities were chosen to replace original ReLUs throughout the network. [[75,76]](https://paperpile.com/c/v2Np13/UfgJw+LESNO) |

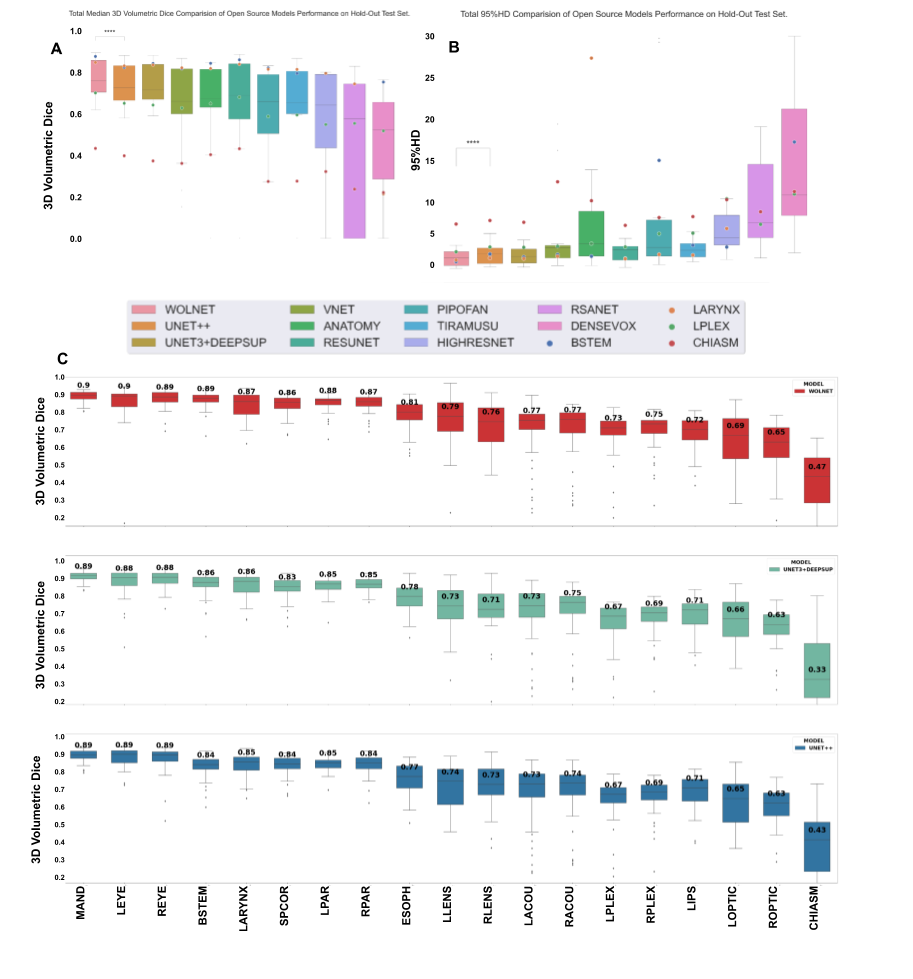

###### **Supplementary Figure 5: Quantitative performance of 11 open-source networks**

Ranking quantitative performance (DICE) of 3 best performing models for 19 OARs. Plotted are the distributions of volumetric dice values for the top performing models when applied to our test set, notice WOLNET is the single 3D CNN that produced superior segmentations for every OAR in our analysis.

#### Performance Evaluation After Model Fine-Tuning

Mandible (0.92±0.03), Eyes (0.88 ± 0.04), and Spinal Cord ( 0.85± 0.05) received the highest mean DICE, while Chiasm (0.41 ± 0.18), Brachial plexus (0.70 ± 0.12), and Optic Nerves (0.71 ± 0.10) received the lowest mean DICE respectively. Chiasm and the Acoustics (L/R) had significantly higher variance in DICE than other OARs. Lenses (1.44 ± 0.53), Eyes (1.81 ± 0.75) and Spinal Cord (1.98±0.73) had the lowest HD, whereas Acoustics (3.66 ± 9.93), Brachial Plexus (4.98 ± 7.35) and Chiasm (6.59 ± 4.46) had the highest recorded 95HD respectively. Brachial Plexus (L/R) and Acoustics (R/L) had significantly higher variance in 95HD than other OARs

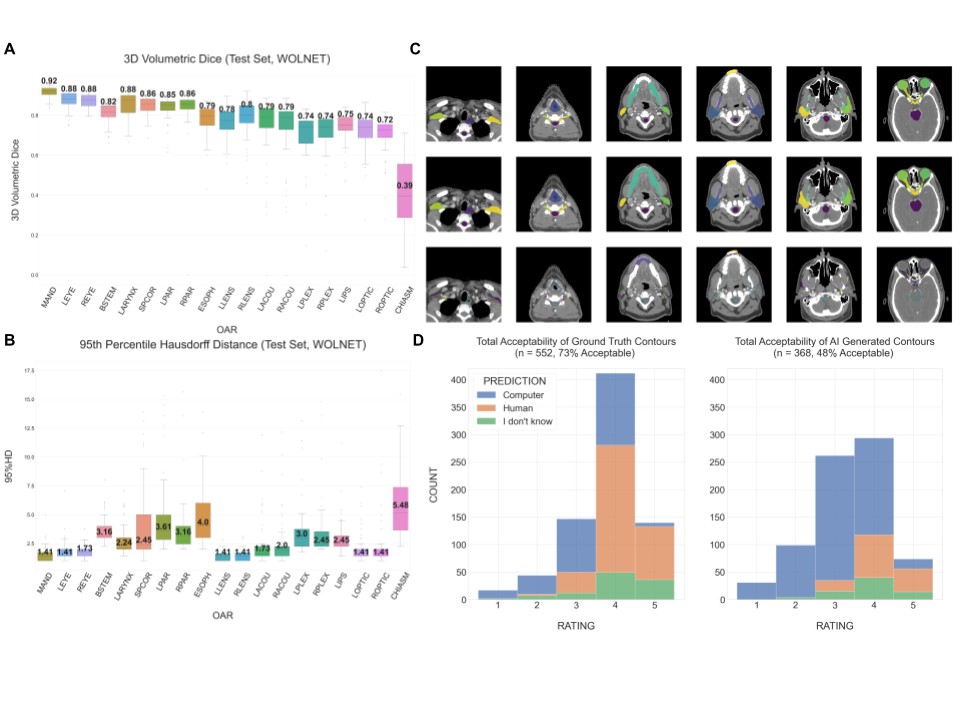

###### **Supplementary Figure 6: Preliminary results for organs-at-risk (OARs) segmentation for re-trained WOLNET**

Preliminary results for organs-at-risk (OARs) segmentation for the best performing network of the 11 re-implemented models (WOLNET) on test set samples in RADCURE. A. 3D Volumetric Dice for each OAR B. Contours from random test set patient C. 95th Percentile Hausdorff Distance for each OAR D. Preliminary results for the clinical acceptability test of the WOLNET predictions using our open-source where 73% of manual contours analysed were considered acceptable, compared with 48% of deep learning contours.

#### **Clinical Evaluation**

To evaluate the clinical acceptability of the OAR contours predicted by the best auto-segmentation model, a subset of 380 contours were reviewed and graded by the oncologists. Results were extracted and Mean Acceptability Rating (MAR) for each OAR was calculated by averaging the ratings obtained for each contour across all observers. The Ground-Truth contours received a global mean acceptability rating (MAR) of 3.81±0.88 and compared with 3.37±0.97 for AI-generated contours (Supplementary Figure 6D). When assessing whether certain OAR categories passed the mean acceptability cutoff of 3.5, 15 Ground-Truth OARs on average were considered clinically acceptable, requiring no edits for planning purposes, compared with 9 AI-generated OARs using the auto-segmentation model. When analysing categories of OARs requiring minor edits for their contours to be accepted into radiation therapy plans (3.0 < MAR < 3.5), 7 AI-generated OARs compared with 4 Ground-Truth OARs met this criteria. The least clinically acceptable Ground-Truth OAR was the chiasm with an MAR of 3.03±1.27, while the most acceptable Ground-Truth OAR was the mandible with an MAR of 4.15±0.89. The least clinically acceptable AI-generated OAR contour was the larynx with an MAR of 2.38±0.90, while the most acceptable AI-generated OAR was the lenses with an MAR of 3.90±0.85

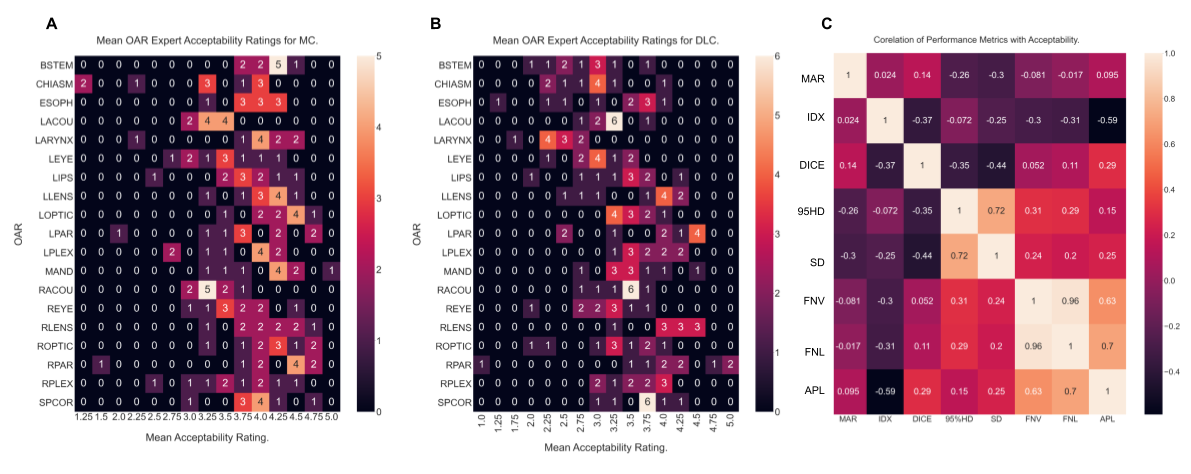

###### **Supplementary Figure 7: Heatmaps of Mean Acceptability Rating Correlation**

Heatmap plots mean acceptability rating correlation with 6 common segmentation metrics. Mean acceptability rating showed significant negative correlation with boundary distance metrics like 95HD and Surface distances. (~-0.26 for 95HD and ~-0.30 for Surface Distance). A less significant positive correlation with DICE was also observed (~0.14).

#### **Model Performance On External Datasets**

The top performing OAR category for the RHN1 dataset when compared against Ground-Truth contours was the mandible with a median DICE of 0.85±0.05 and a median 95HD of 4.0±2.01. In contrast, the least performing OAR categories (with more than one Ground-Truth mask present in the dataset) were the Acoustics, with median DICE of 0.32±0.17 and median 95HD of 7.04±10.35. The HNSCC-3DCT-RT (H3DR) [[46]](https://paperpile.com/c/v2Np13/3OS3A) and StructSeg19 (SS19) [[52]](https://paperpile.com/c/v2Np13/Bzp7J) datasets had 14 out of the 19 RADCURE OARs delineated in their dataset. The top performing OAR categories when compared against Ground-Truth contours in H3DR were the eyeballs (L/R) both achieving median DICE scores of 0.83±0.05 and median 95HD scores of 2.83±0.87 and 2.45±0.64 respectively. The lowest performing OAR in H3DR was the chiasm achieving a median DICE of 0.27±0.21 and a median 95HD of 5.14±3.53. The top performing OAR categories when compared against Ground-Truth contours in SS19 were the parotids (L/R) both achieving median DICE scores of 0.86±0.04 and median 95HD scores of 3.0±1.73 and 3.0±3.0 respectively. The lowest performing OAR in SS19 was also the Chiasm with an average median DICE of 0.33±0.17 and average median 95HD of 5.14±3.53.

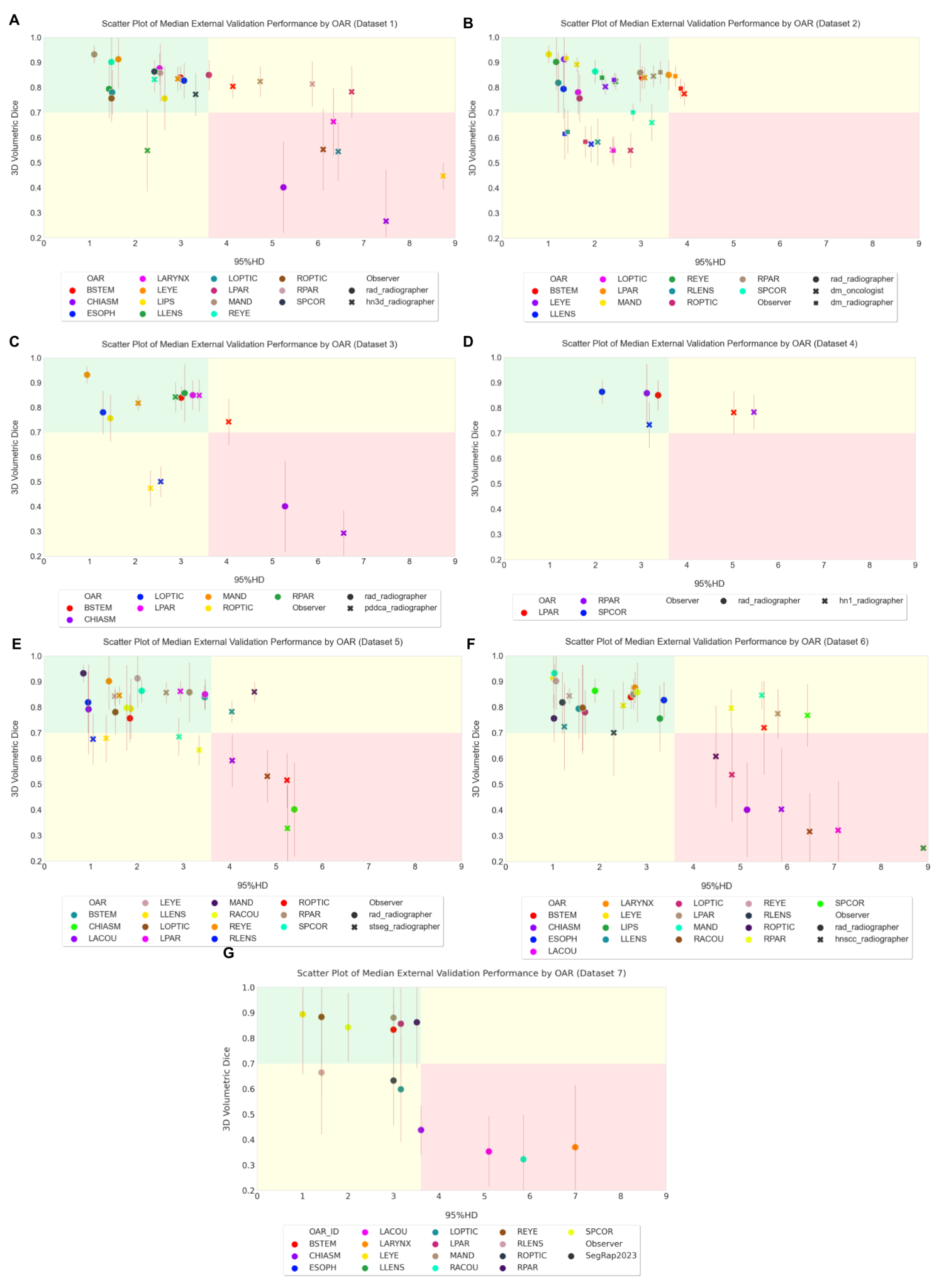

#### Supplementary Figure 8: External Validation Performance of Optimised WOLNET Ensemble across 6 collected external datasets.

Panels A-F Scatter Plots plot median classical performance metrics (DICE v. 95HD) for each OAR category with ground truth labels present in the dataset and compare it against the performance of the ensemble on Radcure's independent test set. OAR(s) that lie in the green shaded region for any given plot, can be considered to lie within the acceptable range of 'interobserver' variability. OAR(s) that lie in the yellow shaded region for any given plot, may require minor edits to lie within the acceptable range of 'interobserver' variability. OAR(s) that lie in the red shaded region for any given plot, may require major edits to lie within the acceptable range of 'interobserver' variability. Panel A: Dataset 1: TCIA-HNSCC-3DCT-RT (n=83) B: Dataset 2: Deepmind (n=35) C: Dataset 3: MACCAI’15 - PDDCA (n=48) D: Dataset 4: TCIA-HN1-RADIOMICS (n=119) E: Dataset 5: MACCAI’19 - StructSeg19 (n=50) F: Dataset 6: TCIA-HNSCC (n=132) G Dataset 7: SegRap2023 (n=120)

| Supplementary Table 4: Median 3D Volumetric DICE Performance of Optimised WOLNET Ensemble on External Data  *** two contours (L/R) mandible were used as single GT mask**  **** Results shown are ensemble application without fine tuning or post processing** | | | | | | | | | |
| --- | --- | --- | --- | --- | --- | --- | --- | --- | --- |
| **OAR** | **RADCURE (n=59)** | **HNSCC-3DCT-RT (n=94)** | **Deepmind - Onc (n=35)** | **Deepmind- Rad (n=35)** | **PDDCA (n=48)** | **TCIA-HNSCC (n=215)** | **STRUCTSEG-19 (n=50)** | **Radiomic -HN1 (n=137)** | **SegRap2023**  **(n=120)** |
| **BSTEM** | 0.84±0.05 | 0.8±0.05 | 0.77±0.05 | 0.8±0.04 | 0.74±0.09 |  | 0.78±0.04 | 0.82±0.08 | 0.83±0.10 |
| **CHIASM** | 0.4±0.18 | 0.27±0.21 |  |  | 0.29±0.09 |  | 0.33±0.17 | 0.4±0.24 | 0.43±0.10 |
| **ESOPH** | 0.83±0.07 | 0.53±0.21 |  |  |  |  |  | 0.69±0.13 | 0.60±0.21 |
| **LACOU** | 0.79±0.17 | 0.18±0.08 |  |  |  | 0.08±0.08 | 0.59±0.1 | 0.27±0.19 | 0.35±0.14 |
| **LARYNX** | 0.88±0.06 | 0.66±0.14 |  |  |  |  |  | 0.49±0.23 | 0.37±0.25 |
| **LEYE** | 0.91±0.12 | 0.83±0.05 | 0.8±0.03 | 0.83±0.03 |  |  | 0.84±0.04 | 0.81±0.09 | 0.89±0.24 |
| **LIPS** | 0.76±0.13 | 0.45±0.05 |  |  |  |  |  | 0.25±0.00 |  |
| **LLENS** | 0.79±0.12 | 0.55±0.16 | 0.57±0.07 | 0.62±0.1 |  |  | 0.68±0.09 | 0.72±0.17 | 0.66±0.08 |
| **LOPTIC** | 0.78±0.09 | 0.54±0.12 | 0.55±0.05 | 0.55±0.06 | 0.5±0.06 |  | 0.53±0.1 | 0.54±0.18 | 0.60±0.21 |
| **LPAR** | 0.85±0.06 | 0.78±0.1 | 0.84±0.04 | 0.84±0.04 | 0.85±0.06 | 0.78±0.09 | 0.86±0.04 | 0.77±0.11 | 0.86±0.21 |
| **LPLEX** | 0.72±0.13 |  |  |  |  |  |  |  |  |
| **MAND** | 0.93±0.03 | 0.82±0.06 | 0.89±0.03 | 0.92±0.02 | 0.82±0.03 |  | 0.86±0.04* | 0.85±0.07 | 0.88±0.16* |
| **RACOU** | 0.8±0.17 | 0.17±0.09 |  |  |  | 0.08±0.04 | 0.63±0.06 | 0.31±0.16 | 0.31±0.17 |
| **REYE** | 0.9±0.11 | 0.83±0.05 | 0.82±0.03 | 0.84±0.03 |  |  | 0.85±0.03 | 0.84±0.03 | 0.88±0.18 |
| **RLENS** | 0.82±0.12 |  | 0.58±0.09 | 0.62±0.09 |  |  | 0.68±0.1 | 0.7±0.17 | 0.66±0.24 |
| **ROPTIC** | 0.76±0.09 | 0.55±0.16 | 0.55±0.07 | 0.58±0.06 | 0.47±0.07 |  | 0.51±0.11 | 0.61±0.2 | 0.63±0.18 |
| **RPAR** | 0.86±0.12 | 0.81±0.09 | 0.85±0.04 | 0.86±0.04 | 0.84±0.06 | 0.78±0.07 | 0.86±0.04 | 0.79±0.1 | 0.86±0.18 |
| **RPLEX** | 0.72±0.14 |  |  |  |  |  |  |  |  |
| **SPCOR** | 0.86±0.05 | 0.78±0.09 | 0.66±0.07 | 0.7±0.04 |  | 0.73±0.09 | 0.68±0.07 | 0.81±0.07 | 0.84±0.13 |

| Supplementary Table 5: Median 95%HD Performance of Optimised WOLNET Ensemble on External Data  *** two contours (L/R) mandible were used as single GT mask**  **** Results shown are ensemble application without fine tuning or post processing** | | | | | | | | | |
| --- | --- | --- | --- | --- | --- | --- | --- | --- | --- |
| **OAR** | **RADCURE (n=59)** | **HNSCC-3DCT-RT (n=94)** | **Deepmind - Onc (n=35)** | **Deepmind- Rad (n=35)** | **PDDCA (n=48)** | **TCIA-HNSCC (n=215)** | **STRUCTSEG-19 (n=50)** | **Radiomics-HN1 (n=137)** | **SegRap23 (n=120)** |
| BSTEM | 3.0±0.88 | 4.12±1.77 | 4.12±0.73 | 3.74±0.68 | 4.0±2.16 |  | 3.74±0.85 | 4.12±2.6 | 3.00±0.79 |
| CHIASM | 5.1±3.94 | 7.26±3.75 |  |  | 6.4±2.62 |  | 5.14±3.53 | 5.62±5.7 | 3.61±1.14 |
| ESOPH | 3.16±5.48 | 14.21±9.13 |  |  |  |  |  | 5.74±3.0 | 14.16±1.12 |
| LACOU | 1.57±9.79 | 9.13±1.93 |  |  |  | 10.1±7.65 | 3.74±1.14 | 7.07±18.69 | 5.10±9.29 |
| LARYNX | 2.24±1.38 | 6.0±2.71 |  |  |  |  |  | 6.66±10.41 | 7.00±0.65 |
| LEYE | 1.41±2.98 | 2.83±0.87 | 2.24±0.16 | 2.24±0.15 |  |  | 1.41±0.39 | 2.45±18.27 | 1.00±0.60 |
| LIPS | 2.45±1.21 | 8.6±2.59 |  |  |  |  |  | 8.6¬±0.00 |  |
| LLENS | 1.41±0.33 | 2.13±0.92 | 2.0±0.31 | 1.65±0.55 |  |  | 1.41±0.37 | 1.41±13.64 | 1.41±17.47 |
| LOPTIC | 1.41±1.29 | 6.48±5.28 | 2.24±1.53 | 2.24±1.88 | 2.63±4.44 |  | 5.05±2.19 | 4.24±16.6 | 3.16±0.50 |
| LPAR | 3.32±3.44 | 6.4±4.41 | 3.32±2.77 | 3.74±5.17 | 3.16±1.72 | 4.9±3.52 | 3.0±1.73 | 5.79±4.64 | 3.16±0.46 |
| LPLEX | 4.0±13.93 |  |  |  |  |  |  |  |  |
| MAND | 1.0±12.19 | 4.29±4.47 | 1.41±0.32 | 1.0±0.42 | 2.0±23.97 |  | 4.3±14.11* | 4.0±2.01 | 3.00±13.3 |
| RACOU | 1.73±9.95 | 9.95±3.98 |  |  |  | 11.8±21.7 | 3.73±0.97 | 7.0±2.87 | 5.86±4.75 |
| REYE | 1.41±2.97 | 2.45±0.64 | 2.24±0.14 | 2.0±0.13 |  |  | 1.41±0.3 | 2.12±0.21 | 1.41±2.41 |
| RLENS | 1.41±0.45 |  | 1.95±0.31 | 1.41±0.44 |  |  | 1.41±0.41 | 1.87±14.37 | 1.41±5.90 |
| ROPTIC | 1.41±1.6 | 6.04±5.14 | 2.24±0.42 | 2.0±1.34 | 2.24±1.65 |  | 5.03±2.68 | 4.61±14.17 | 3.00±2.46 |
| RPAR | 3.0±4.29 | 5.74±4.86 | 3.16±2.36 | 3.16±2.73 | 3.0±1.66 | 5.39±4.75 | 3.0±3.04 | 5.16±4.25 | 3.51±3.68 |
| RPLEX | 4.0±13.8 |  |  |  |  |  |  |  |  |
| SPCOR | 2.0±1.3 | 3.0±4.35 | 3.16±1.27 | 3.0±1.41 |  | 3.0±12.3 | 2.83±0.83 | 2.83±4.74 | 2.00±3.65 |

### **Appendix C: Discussion**

We trained 11 open source segmentation architectures on both internal and external datasets and identified the top-performing 3D UNET network, WOLNET, for the acceptability phase. We fine-tuned the WOLNET model and assessed its segmentation quality using the clinical evaluation toolkit, QUANNOTATE, for blinded acceptability review. Demonstrating model robustness, we tested the WOLNET on seven publicly available external datasets. Our results show that the simple 3D UNET provides the best objective auto-segmentation performance among the tested models.

It is important to note that multiple metrics are required when assessing performance of segmentation methods because metrics can vary dramatically when assessing mistakes in large or small regions of interest. In our experiments, the Chiasm contour, which spans two to three axial slices at the top of a scan, is exponentially smaller in voxel size than the larger organs-at-risk being segmented in our training task. Moreover, a small miss annotated pixel in small OARs like the chiasm class, will have a dramatic negative impact on both average Dice and Hausdorff Distance [[77,78]](https://paperpile.com/c/v2Np13/SO9Q8+lJAe0). The chiasm class has much larger human inter-observability than larger OAR(s) so, in this case, it would be better to assess the model’s performance on general region localization, than penalising the overall model for in-accurately predicting individual pixels.

While there currently is no global consensus on interpreting volumetric similarity metrics, studies have suggested that any overlap index ≥ 0.7 could be considered to have “good agreement” [[79]](https://paperpile.com/c/v2Np13/1Yicm). Agreement of any volumetric similarity metric will change based on experience level of the observers, the difficulty of the resulting segmentation task and the modality used for contouring. Previous studies sought to define mean overlap indices between three expert radiation oncologists, each with more than 10 years of treatment experience, for GTV and various OAR contours [[80]](https://paperpile.com/c/v2Np13/tyTLX). Gudi S et. al found the average interobserver dice similarity coefficient of GTV contour on simulation CT(s) to be 0.57±12, therefore a segmentation network would have to “maintain or exceed'' this rating for the GTV contours it produces to be within expert interobserver variability. The study also recorded average interobserver dice values for Right Parotid (0.77±0.04), Left Parotid (0.78±0.04), Right Cochlea (We have this as ‘Right Acoustic’) (0.07±0.08), Left Cochlea (We have this denoted as ‘Right Acoustic’) (.10±0.07) and spinal cord (0.78±0.03) where a similar conclusion can be made. Conducting external validation experiments on multiple datasets without fine tuning to external data was essential to assess the direct generalizability of the network selected for acceptability assessment. Agreement of any volumetric similarity metric will change based on experience level of the observers, the difficulty of the resulting segmentation task and the modality used for contouring.

When comparing these inter observability thresholds for select OARs to results generated by our generalizability assessment, regardless of the dataset, the WOLNET model performed well at segmenting large OARs like the Mandible and Brain Stem and OARs with homogeneous structures like the Eyeballs and Parotids. When comparing these results to metrics obtained when assessing OAR contouring from different human observers, we achieve or beat out human standards of variability. For example, for the parotid contour the WOLNET model met or superseded current interobserver variability 3D DICE thresholds (0.77±0.04) for the parotids across all datasets tested. Similarly, when comparing our results to the human interobserver variability of the spinal cord contour (0.78±0.03) [[80]](https://paperpile.com/c/v2Np13/tyTLX), we found that this threshold was upheld or exceeded for 3 out of the 6 datasets tested. Adequate performance across most OARs categories was also achieved across the different datasets except the Acoustics. This was expected as there is very large variability in ground truth contouring of this select OAR among clinicians, where current inter observability thresholds for 3D DICE are very low (average scores ranging from 0.07-0.10) [[80]](https://paperpile.com/c/v2Np13/tyTLX).

[30]	Jun Ma. SegLoss: A collection of loss functions for medical image segmentation. Github; n.d.

[31]	Liu L. RAdam. Github; n.d.

[32]	[Liu L, Jiang H, He P, Chen W, Liu X, Gao J, et al. On the Variance of the Adaptive Learning Rate and Beyond. arXiv [csLG] 2019.](http://paperpile.com/b/v2Np13/4oTcT)

[33]	Beare R, Lowekamp B, Yaniv Z. Image segmentation, registration and characterization in R with SimpleITK. J Stat Softw 2018;86. https://doi.org/[10.18637/jss.v086.i08](http://dx.doi.org/10.18637/jss.v086.i08).

[34]	Eric Kerfoot, Raghav Mi, Tom Vercauteren, Wenqi Li. MONAI: AI Toolkit for Healthcare Imaging Release 0.5.3. Github; n.d.

[35]	[Zhang C, Bengio S, Hardt M, Recht B, Vinyals O. Understanding deep learning requires rethinking generalization. arXiv [csLG] 2016.](http://paperpile.com/b/v2Np13/dKrEz)

[36]	[Thambawita V, Hicks SA, Halvorsen P, Riegler MA. DivergentNets: Medical Image Segmentation by Network Ensemble. arXiv [eessIV] 2021.](http://paperpile.com/b/v2Np13/sKNq8)

[37]	Li R, Auer D, Wagner C, Chen X. A Generic Ensemble Based Deep Convolutional Neural Network for Semi-Supervised Medical Image Segmentation. 2020 IEEE 17th International Symposium on Biomedical Imaging (ISBI) 2020. https://doi.org/[10.1109/isbi45749.2020.9098568](http://dx.doi.org/10.1109/isbi45749.2020.9098568).

[38]	Kamraoui RA, Ta V-T, Tourdias T, Mansencal B, Manjon JV, Coup P. DeepLesionBrain: Towards a broader deep-learning generalization for multiple sclerosis lesion segmentation. Med Image Anal 2022;76:102312.

[39]	Tomara NK, Ibtehazc N, Jhaa D, Halvorsena P, Alid S. Improving Generalizability in Polyp Segmentation using Ensemble Convolutional Neural Network 2021.

[58]	Wolny A. pytorch-3dunet: 3D U-Net model for volumetric semantic segmentation written in pytorch. Github; n.d.

[59]	[Lee K, Zung J, Li P, Jain V, Sebastian Seung H. Superhuman Accuracy on the SNEMI3D Connectomics Challenge. arXiv [csCV] 2017.](http://paperpile.com/b/v2Np13/q88Oc)

[60]	[Li W, Wang G, Fidon L, Ourselin S, Jorge Cardoso M, Vercauteren T. On the Compactness, Efficiency, and Representation of 3D Convolutional Networks: Brain Parcellation as a Pretext Task. arXiv [csCV] 2017.](http://paperpile.com/b/v2Np13/QFAZb)

[61]	Nikolas A. HighResNet3D.py at master · black0017/MedicalZooPytorch. Github; n.d.

[62]	Fang X, Yan P. Multi-Organ Segmentation Over Partially Labeled Datasets With Multi-Scale Feature Abstraction. IEEE Trans Med Imaging 2020;39:3619–29.

[63]	PIPO-FAN: PIPO-FAN for multi organ segmentation over partial labeled datasets using pytorch. Github; n.d.

[64]	Huang H, Lin L, Tong R, Hu H, Zhang Q, Iwamoto Y, et al. UNet 3+: A Full-Scale Connected UNet for Medical Image Segmentation. ICASSP 2020 - 2020 IEEE International Conference on Acoustics, Speech and Signal Processing (ICASSP), 2020, p. 1055–9.

[65]	UNet-Version. Github; n.d.

[66]	Zhou Z, Siddiquee MMR, Tajbakhsh N, Liang J. UNet++: A Nested U-Net Architecture for Medical Image Segmentation. Deep Learn Med Image Anal Multimodal Learn Clin Decis Support (2018) 2018;11045:3–11.

[67]	UNet-Version. Github; n.d.

[68]	Zhu W. AnatomyNet-for-anatomical-segmentation: AnatomyNet: Deep 3D Squeeze-and-excitation U-Nets for fast and fully automated whole-volume anatomical segmentation. Github; n.d.

[69]	[Yu L, Cheng J-Z, Dou Q, Yang X, Chen H, Qin J, et al. Automatic 3D Cardiovascular MR Segmentation with Densely-Connected Volumetric ConvNets. arXiv [csCV] 2017.](http://paperpile.com/b/v2Np13/vzQuz)

[70]	Nikolas A. DenseVoxelNet.py at master · black0017/MedicalZooPytorch. Github; n.d.

[71]	Fortuner B. pytorch_tiramisu: FC-DenseNet in PyTorch for Semantic Segmentation. Github; n.d.

[72]	[Jégou S, Drozdzal M, Vazquez D, Romero A, Bengio Y. The One Hundred Layers Tiramisu: Fully Convolutional DenseNets for Semantic Segmentation. arXiv [csCV] 2016.](http://paperpile.com/b/v2Np13/SA3QS)

[73]	Zhang H, Zhang J, Zhang Q, Kim J, Zhang S, Gauthier SA, et al. RSANet: Recurrent Slice-Wise Attention Network for Multiple Sclerosis Lesion Segmentation. Medical Image Computing and Computer Assisted Intervention – MICCAI 2019, Springer International Publishing; 2019, p. 411–9.

[74]	tinymilky. RSANet: RSANet: Recurrent Slice-wise Attention Network for Multiple Sclerosis Lesion Segmentation (MICCAI 2019). Github; n.d.

[75]	[Milletari F, Navab N, Ahmadi S-A. V-Net: Fully Convolutional Neural Networks for Volumetric Medical Image Segmentation. arXiv [csCV] 2016.](http://paperpile.com/b/v2Np13/UfgJw)

[76]	Macy M. vnet.pytorch: A PyTorch implementation for V-Net: Fully Convolutional Neural Networks for Volumetric Medical Image Segmentation. Github; n.d.

[77]	[Maier-Hein L, Reinke A, Godau P, Tizabi MD, Büttner F, Christodoulou E, et al. Metrics reloaded: Pitfalls and recommendations for image analysis validation. arXiv [csCV] 2022.](http://paperpile.com/b/v2Np13/SO9Q8)

[78]	Reinke A, Tizabi MD, Baumgartner M, Eisenmann M, Heckmann-Nötzel D, Kavur AE, et al. Understanding metric-related pitfalls in image analysis validation. ArXiv 2023. https://doi.org/[10.3115/1072064.1072067](http://dx.doi.org/10.3115/1072064.1072067).

[79]	Vinod SK, Min M, Jameson MG, Holloway LC. A review of interventions to reduce inter-observer variability in volume delineation in radiation oncology. J Med Imaging Radiat Oncol 2016;60:393–406.

[80]	Gudi S, Ghosh-Laskar S, Agarwal JP, Chaudhari S, Rangarajan V, Paul SN, et al. Interobserver Variability in the Delineation of Gross Tumour Volume and Specified Organs-at-risk During IMRT for Head and Neck Cancers and the Impact of FDG-PET/CT on Such Variability at the Primary Site. Journal of Medical Imaging and Radiation Sciences 2017;48:184–92. https://doi.org/[10.1016/j.jmir.2016.11.003](http://dx.doi.org/10.1016/j.jmir.2016.11.003).
